# Exploring associations between trust in doctors and the likelihood of accessing healthcare for anxiety, depression and menstrual problems

**DOI:** 10.64898/2026.09.30.26364328

**Authors:** Diya Hoque, Robyn E Wootton, Claire M A Haworth, Kayleigh E Easey

## Abstract

**Background:** Anxiety, depression and menstrual problems are conditions that significantly impact an individual’s health and wellbeing yet, barriers to healthcare can prevent access to appropriate medical care. Trust in doctors is a notable factor as previous studies have shown that it affects engagement with healthcare services and has declined in recent years.

**Methods:** Using the Avon Longitudinal Study of Parents and Children, we investigated associations between trust in doctors and healthcare access at two timepoints, for anxiety, depression (mothers and partners) and menstrual problems (mothers only).

**Results:** Mothers reporting greater trust in doctors were more likely to access care for anxiety (n=1849, adjusted odds ratio (AOR) 1.11, 95% confidence interval (95% CI) 1.08-1.14), depression (n=2411, AOR 1.10, 95% CI 1.07-1.12), and menstrual problems (n=1723, AOR 1.04 95% CI 1.01-1.06). If mothers and partners endorsed “the doctor is always helpful”, they were more likely to access care for depression (mothers: n=2005, AOR 1.30, 95% CI 1.16-1.46; partners: n=580, AOR 1.46, 95% CI 1.05-2.03) and for anxiety (mothers: n=1892, AOR 1.35, 95% CI 1.19 to 1.54; partners: n=734, AOR 1.40, 95% CI 1.02-1.91).

**Implications:** This study supports association between trust in doctors and healthcare access for anxiety, depression and menstrual problems, particularly amongst mothers. This contributes to previous literature that suggests that trust remains an important factor in understanding treatment gaps, and that researchers and clinicians must work to improve public trust in healthcare systems.

**Lay Summary:** The number of people experiencing anxiety, depression and menstrual problems is rising. All three conditions can dramatically affect an individual’s quality of life however, many people report feeling unable to seek out professional help. Known as the treatment gap, the difference between those who require medical care and those who receive it, is driven by several factors, highlighting the need to understand the factors that determine whether people access healthcare. Trust in doctors is a notable factor, as previous studies have shown that it affects healthcare access and has declined in recent years. As such, this study seeks to understand whether there is a relationship between trust in doctors and healthcare access for anxiety, depression and menstrual problems.

This study used data from the Avon Longitudinal Study of Parents and Children (ALSPAC), using data from two different timepoints, to assess this relationship amongst mothers and their partners. We investigated this relationship in mothers for anxiety, depression and menstrual problems and in partners for anxiety and depression only.

Results from this study suggest that this relationship was evident in mothers for all three conditions, when their children were 21 months old. There was also some evidence of the relationship between trust in doctors and healthcare access for anxiety and depression, when their child was 6 years old. Partners also showed some evidence of this relationship for anxiety and depression when their child was 6 years old. This work contributes to previous research showing that trust in doctors may be an important factor in determining whether people access healthcare for these conditions and suggests that researchers and doctors should work to improve public trust in healthcare systems.

## Introduction

The prevalence of poor mental health is increasing, with more than 1 billion people worldwide reporting to be living with a mental health condition (1). Anxiety and depression, known as common mental disorders (CMDs), both sit within the top ten causes of disability, with an estimated 4.8% of the global population experiencing an anxiety disorder (2) and 5.7% experiencing a depressive disorder (3). Both conditions can impact daily functioning and overall quality of life (4). Similarly, menstrual disorders can impede daily life, with people experiencing menstrual disorders reporting lower quality of life scores in most domains compared to their counterparts (5). Menstrual disorders are defined as the interruption of the monthly cyclic order and/or some physical dysfunction related to the menstrual cycle (6). Serving as an umbrella term for several conditions, symptoms seen from menstrual disorders include, unusually heavy or light bleeding (7), missed or irregular periods (8) and severe pain (9). Literature on the prevalence of menstrual disorders ranges wildly, from 3% to 87%, depending on the geographic region and methodology used (10–13). This lack of standardisation highlights a challenge in the field, when attempting to understand help-seeking behaviours for menstrual disorders. Both CMDs and menstrual disorders are conditions where individuals often feel unable to seek out professional help (14–16). Consequently, these conditions exhibit large treatment gaps, driven by both structural and individual factors, (15–17), highlighting the need to understand the factors that determine if and why people access healthcare.

A lack of engagement with health services affects an individual’s ability to obtain diagnoses, treatment, and information to support their health (18). For individuals experiencing CMDs and menstrual disorders, many people do not access healthcare upon initial presentation of symptoms. Evidence suggests that, globally, only 36.3% of individuals with symptoms of anxiety receive any sort of treatment (19). Similar was found globally for depression, with only 34.8% of people with symptoms receiving treatment (20). There has not been a synthesis of global evidence on the treatment gap for menstrual problems, however, Uchibori and colleagues (21) showed that 50% of their Japanese cohort recognised a need to access healthcare for menstrual or menopausal symptoms, and of those, only 77% sought care for their symptoms. To improve the treatment gap for these conditions, it is necessary to understand what prevents healthcare access.

The gap between those who need medical care and those who receive it (22) is driven by social, economic, environmental and structural factors (23). One factor that has been explored is the role of trust in physicians and healthcare systems. Trust is one of the fundamental principles of the patient-physician relationship (24) and has been conceptualised in several ways. The overarching theme of these definitions relates to a set of beliefs or expectations patients’ have about how physicians will behave (24), these can be split into concepts of public trust, and interpersonal trust. Interpersonal trust refers to the trust built on the individual relationship between the patient and the physician, developed through repeated interactions (25). Public trust relates to trust in collective institutions and entities (i.e. physicians in general), and is widely influenced by the media and general social confidence (26). Interpersonal trust and public trust are closely linked, with public trust and an individual’s past experiences shaping interpersonal trust (27).

A decline of system-level trust in physicians has been widely observed (28–30), with interpersonal trust likely to follow. This is concerning as a lack of trust in healthcare providers and systems has been associated with lower adherence to treatment, engagement in risky behaviours and poor relationships between patients and physicians (30). Further, greater levels of trust have been associated with fewer reported symptoms and greater satisfaction with care (31). Trust has also been found to affect engagement with health services. Ewald and colleagues (32) have shown that respondents with medical conditions were less likely to delay their medical care if they trusted doctors.

The relationship between trust in physicians and healthcare access is affected by multiple demographic characteristics, such as gender. Adherence to normalised gender roles has been reported to affect healthcare access in several ways. Specifically, women have reported greater healthcare use for anxiety and depression (33), with men being more likely to hold negative attitudes to the use of mental health services (34). However, women have often reported avoiding healthcare for menstrual problems as menstrual pain has been perceived as a standard part of womanhood (15). There is less evidence available to suggest that gender affects patient trust, however, Kim and colleagues (35) reported that men were 1.3 times more likely to trust physicians compared to women. Further investigation is therefore needed to explore whether trust facilitates healthcare access across genders.

The exploration into the role of trust on help-seeking behaviours is particularly important for recent parents, among whom mental disorders are widely considered to be underdiagnosed (36). Parents in the postpartum period are especially vulnerable to several mental disorders due to rapid and often overwhelming biological, social and emotional changes (37). Postpartum depression (PPD) is thought to affect 10-17% of mothers (38,39), however, it is estimated that 50% of mothers with PPD go undiagnosed (40). Incidence of paternal PPD varies widely, with estimates ranging from 4% to 25%, within the first twelve months postpartum (41,42). Studies suggest that paternal PPD is frequently underreported, which may reflect uncertainty in prevalence estimates (43). The estimated global prevalence of postpartum anxiety (PPA) in mothers is between 12% and 27% (44,45) and in fathers it is between 5-15% (46), though it is unclear how many people go undiagnosed with PPA. Understanding the factors that impact help-seeking behaviours of mothers and fathers is especially important to prevent maternal and paternal morbidity, both of which have been associated with significant, long-term mental health effects for both parents and their children (47–49).

This study will therefore explore the association between trust in physicians and healthcare access for depression, anxiety and menstrual problems, to understand the relationship between trust and help-seeking for CMDs and menstrual disorders, amongst mothers and their partners. This study will also explore whether this association remains consistent across mothers and partners.

## Methods

### Sample

Women who were pregnant in the former Avon County, UK, and were expected to give birth between 1st April 1991 and 31st December 1992 were eligible for the ALSPAC study (50). 20,248 pregnancies have been identified as eligible and 14,541 pregnancies were initially recruited, giving a total of 14,676 foetuses, resulting in 14,062 live births and 13,988 children who were alive at the age of 1 (50). Of the original 14,541 initial pregnancies, 338 were from a woman who had already enrolled with a previous pregnancy, meaning 14,203 unique mothers were initially enrolled onto the study (51). When children were 7, an attempt to bolster the initial sample with eligible cases who had failed to join the study initially. Additional participants were recruited and added to the cohort following the initial recruitment period, when the children were of school-age. As a result of additional recruitment, a further 630 women who did not originally enrol have provided data since their child was 7 years old. This provides a total of 14,833 unique women (G0 mothers) enrolled in ALSPAC as of September 2021 (51). G0 mothers were asked to recruit their partners into ALSPAC (G0 partners) and in most cases this was the biological father or father figure of the children (52). ALSPAC has been in contact with 12,113 G0 partners (52). 3,807 G0 partners are currently enrolled (52). Data was routinely collected through postal questionnaires, with the completion of a questionnaire, either on paper or online, was considered to be written consent from participants to use their data for research purposes. Please note that the study website contains details of all the data that is available through a fully searchable data dictionary and variable search tool (http://www.bristol.ac.uk/alspac/researchers/our-data/).

### Measures

Data on the main explanatory variables and outcomes of interest were taken at 2 timepoints; at 21 months postpartum, study mothers were asked questions regarding their trust in doctors and whether they sought healthcare between 8 and 21 months postpartum for anxiety, depression and/or menstrual problems. Study mothers and their partners were asked separately, when their child was 6 years old, about their trust in doctors and whether they had sought healthcare for anxiety and/or depression between their child’s fifth and sixth birthday. Mothers aged 45 years and above at childbirth were excluded as it has been well documented that their experiences of healthcare are especially unique due to often needing more specialised care and more frequent healthcare visits (53).

### Exposures

Trust in doctors was measured differently at the two timepoints: at 21 months postpartum, mothers were asked about their feelings towards their doctor. Mothers were asked if they thought their doctor was “supportive”, “sympathetic”, “interested”, “helpful”, “easy to talk to” and “prepared to give mum time”. Response categories were “never”, “sometimes”, “usually”, “always” within a numerical scale (1-4).

A correlation matrix indicated that these variables were highly correlated (r<0.77) and so were combined into a composite variable, with values ranging from 4 to 24 (Figure S1.).

At 6 years after childbirth, mothers and partners were individually asked 3 questions relating to their trust in doctors: “I don’t have any confidence in the NHS”, “I know that if my child was very ill, a doctor would come quickly” and “the doctor in the clinic is always helpful”. Response options were “I never feel this way”, “this is sometimes how I feel”, “this is often how I feel” and “this is exactly how I feel”, within a numerical scale (1-4). Correlation matrices indicated low correlation scores between these variables Figure S2 and S3.), so these variables were kept separate.

### Outcomes

At 21 months postpartum, mothers were asked if they had experienced anxiety, depression or menstrual problems since their toddler was 8 months old. Response options were “yes and consulted a doctor”, “yes but did not consult a doctor” and “no”. As this research aimed to explore the association between trust in doctors and the likelihood of seeking care, those who had answered “no” to experiencing a condition, were excluded and the remaining responses were dichotomised, where 0= those who experienced the condition but did not consult a doctor and 1= those who experienced the condition and did consult a doctor. At 6 years postpartum, mothers and partners were separately asked if they had experienced anxiety or depression since their child was 5 years old. Responses were again binarised, the same as for 21 months.

### Confounders

We adjusted for confounders that have been shown to be associated with both trust in doctors and healthcare access: age (54), marital status (55), home ownership (56), parity (57), educational attainment (58), religion (59), socioeconomic position (60), self-rated health (61), social support (62), significant life events (63), crown-crisp anxiety and depression scores (64) and a recent doctor change (65). The measures used for each of these confounders are described in the Supplementary Note.

## Statistical analyses

Analyses were performed using RStudio version 4.5.1. Logistic regressions explored associations between trust in doctors and the likelihood of seeking care for anxiety, depression, and (mothers only) menstrual problems. Comparisons were made between those who experienced a condition and consulted a doctor for it to those who experienced the same condition but did not consult a doctor.

Unadjusted models were compared to adjusted models to explore the impact of confounding on the association of interest. Model 1 (baseline demographics) was adjusted for age, socioeconomic position, educational attainment, parity, home ownership status, marital status and religion. Model 2 (relationship with healthcare) was further adjusted for recent doctor changes and evaluation of own health and model 3 (social networks) was further adjusted for social support score and weighted life events score.

The sample size decreased significantly following each adjustment and so analyses were first conducted using the maximum available sample for each specific model, regardless of missingness of the other variables (Table S9, S12, S15). To assesses the robustness of these findings against missing data, then the analysis was repeated including only participants with complete data. Complete case analysis, the method of only including data records without missing values for any variable, was used in the first instance to remove missing data (66). However, the cumulative effect of missing data in multiple variables often leads to the exclusion of a significant proportion of participants in the original sample, which affects both precision, power and possibly bias depending on patterns of missingness (67). The missingness of each variable in the dataset varied from 0.16% to 51.56%. As such, multiple imputation was also conducted to compare with results from the complete case analysis (Table S10, S13, S16). Multiple imputation generates plausible values for missing data derived from distributions of and relationships among observed variables in the dataset (68). Multiple imputation by chained equations (MICE) was conducted in R, generating 50 imputed datasets, each with 20 cycles. The generation of multiple datasets provides a quantification of the uncertainty in estimating what the missing values might be, therefore avoiding false precision (68). Results generated from imputed data are given in the main text and results from the original and complete case dataset are included in the supporting information.

## Results

### 21-month timepoint

At 21 months, 18% of mothers reported experiencing anxiety (1849 out of 9915), 24% reported experiencing depression (2411 out of 9960) and 17% reported experiencing menstrual problems (1723 out of 10,007). Of those experiencing these conditions, 26% of mothers (489 out of 1849) consulted a doctor for anxiety, 33% (786 out of 2411) for depression and 62% (1068 out of 1723) for menstrual problems. Additional descriptive statistics are available in the supplementary material (Table S1.).

**Figure 1.**
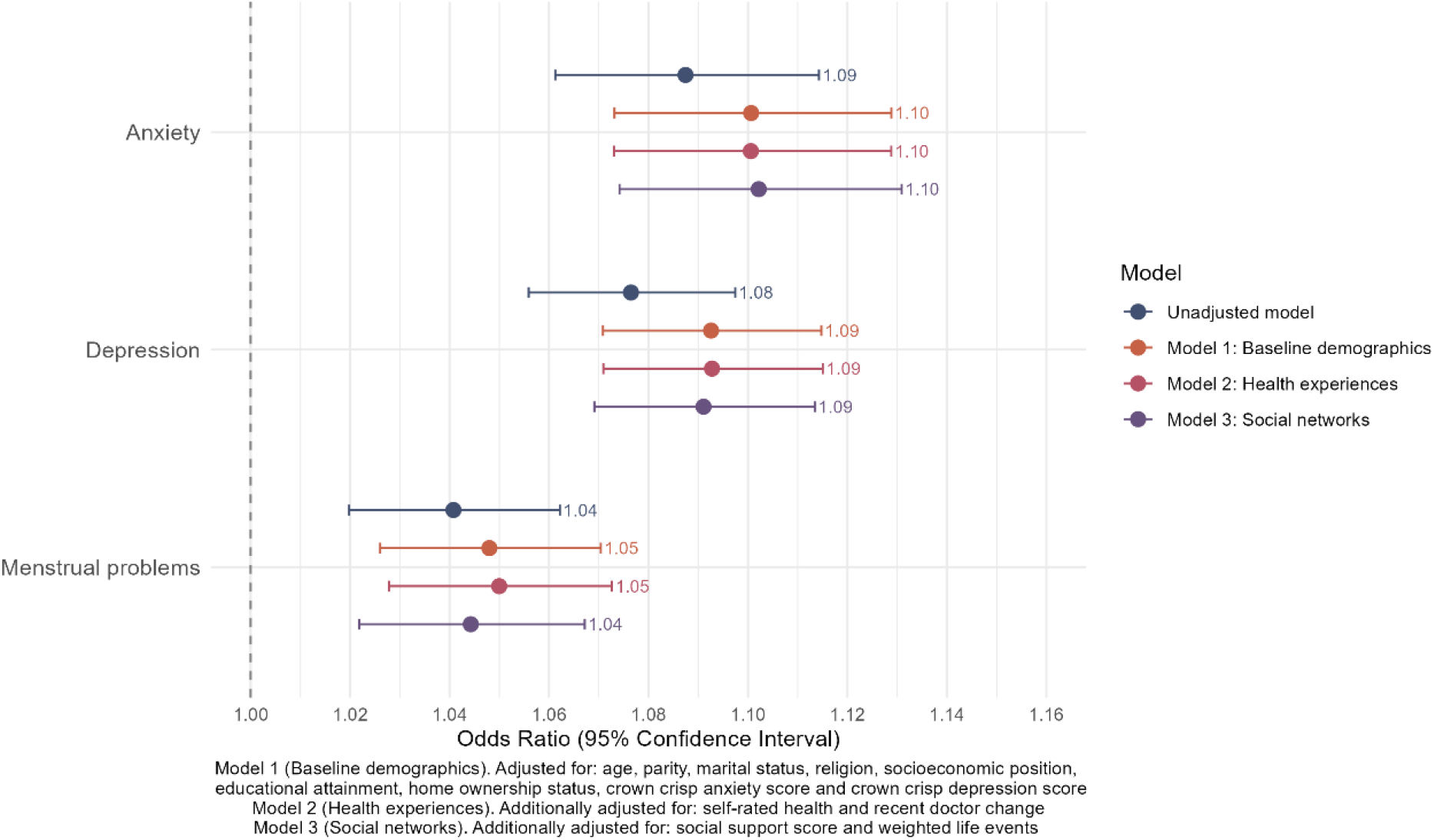
Forest plot of unadjusted and adjusted models exploring associations between trust in doctors and healthcare access for anxiety, depression and menstrual problems at 21 months, using imputed data

Both unadjusted and adjusted models found participants who were more trusting of their doctor, were more likely to consult them for anxiety, depression and menstrual problems. For anxiety, when fully adjusted, the odds ratio of this association was 1.10 (95% CI 1.07-1.13). Similar associations were observed for those experiencing depression, with the fully adjusted odds ratio for this association being 1.09 (95% CI 1.07-1.11). The association between trust in doctors and healthcare access was slightly weaker for those experiencing menstrual problems, compared to anxiety and depression with the adjusted odds ratio for this association being 1.04 (95% CI 1.02-1.07), when fully adjusted. The observed associations for anxiety, depression, and menstrual problems were largely unchanged after adjustment for age, parity, marital status, religion, socioeconomic position, educational attainment, home ownership status, crown-crisp anxiety score, crown-crisp depression score, self-rated health, recent doctor change, social support score, and weighted life events score, with only slight attenuation of the effect estimates.

### 6-year timepoint

Of the 22% of mothers reporting symptoms of anxiety (1892 out of 8435), 31% (594 out of 1892) reported consulting a doctor for anxiety. 23% reported experiencing depression (2005 out of 8439) and 42% (842 out of 2005) consulted a doctor for their symptoms. Out of the 16% of partners that reported experiencing anxiety (725 out of 4368), 16% consulting a doctor for anxiety (120 out of 725). For the 13% of partners experiencing depression symptoms (569 out of 4364), 24% consulted a doctor (138 out of 569). Additional descriptive statistics are available in the supplementary material (Table S2., S3., S4., S5., S6., S7.).

**Figure 2.**
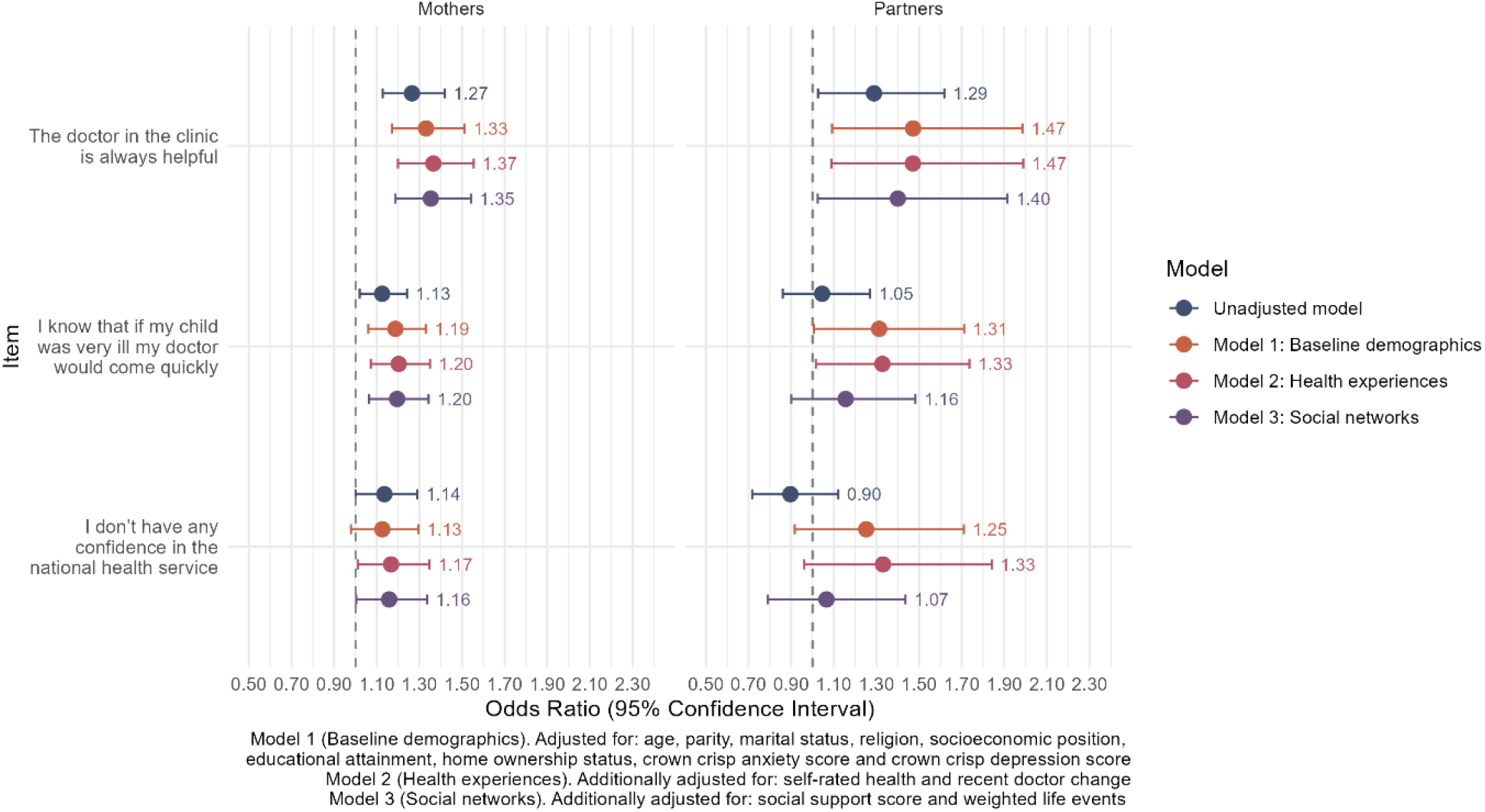
Forest plot of unadjusted and adjusted models exploring associations between trust in doctors and healthcare access for anxiety, at 6 years after baseline, using imputed data

There was mixed evidence of the association between trust in doctors and healthcare access for both mothers and partners, at 6 years. Believing that the doctor in the clinic is always helpful was associated with healthcare access for anxiety for both mothers and partners. In fully adjusted analyses, the effect size was greater in partners (OR 1.40, 95% CI 1.02 – 1.91) compared to mothers (OR 1.35, 95% CI 1.19 – 1.54). Believing a doctor would come quickly if their child was ill was associated with healthcare access for anxiety for mothers across all levels of adjustment but not partners. The effect size for this association, for mothers, was 1.20 (95% CI 1.06 – 1.34) when fully adjusted. The only evidence of this association for partners was when for adjusting for model 2 (health experiences) (OR 1.33 95% CI 1.02 - 1.74). Reporting greater confidence in the NHS did not increase the likelihood of accessing healthcare for anxiety for mothers or partners.

**Figure 3.**
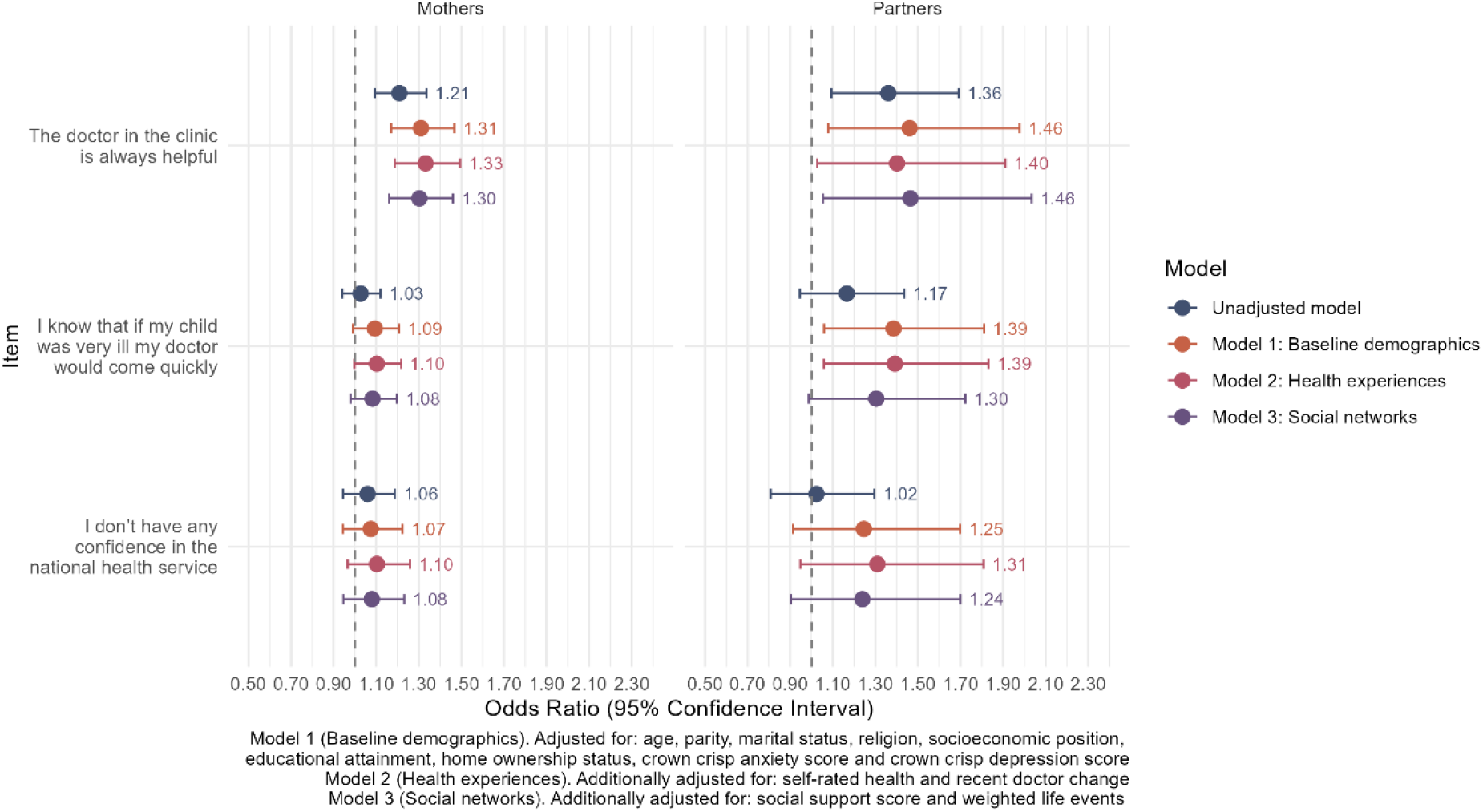
Forest plot of unadjusted and adjusted models exploring associations between trust in doctors and healthcare access for depression, at 6 years after baseline, using imputed data

Believing that the doctor in the clinic is always helpful was associated with healthcare access for depression for both mothers and partners. The fully adjusted odds ratios were 1.30 (95% CI 1.16–1.46) among mothers and 1.46 (95% CI 1.05–2.03) among partners. However, believing that a doctor would come quickly if their child was ill or having confidence in the NHS did not impact healthcare access for depression for both mothers and partners.

## Discussion

This study used the ALSPAC birth cohort to understand the relationship between trust in doctors and healthcare access for anxiety, depression and menstrual problems among mothers and their partners. Overall, respondents who expressed greater levels of trust in doctors were more likely to access healthcare for all three conditions. When index children were 21 months, this association was observed consistently for anxiety, depression and menstrual problems and when index children were 6 years old, both mothers and partners demonstrated some evidence for this association, but not for all items. Differences in these associations at the two timepoints may reflect how trust changes over the course of parenthood. Healthcare contact is more frequent when a child is younger due to milestone screenings and routine immunisations (69). Trust may have been particularly important for mothers, when deciding whether to disclose health concerns to their doctors, for conditions that are especially stigmatised for mothers, such as anxiety and depression (70). Further, by 21 months postpartum, mothers usually experience a return to their regular menstrual cycle, following pregnancy and lactation (71). Trust may influence whether mothers seek out care for menstrual irregularities during this period, as it is closely associated with patients’ perceptions of feeling understood by their healthcare providers (72). Mothers would have to trust that their doctors will acknowledge and not dismiss their concerns around menstrual disorders, which are frequently overlooked by clinicians (15). By 6 years, parents typically attend fewer appointments with their children, reducing their engagement with healthcare services (73). This reduction in healthcare interaction may affect mothers’ trust in doctors and their willingness to access healthcare for their own conditions. Furthermore, differences in associations at 21 months and 6 years may reflect differences in the operationalisation of trust at each questionnaire. At 21 months, variables are specifically and only asking participants about their relationship with their doctor, relating to interpersonal trust. In contrast, variables at 6 years asked participants about their opinions on their own doctor (“the doctor in the clinic is always helpful”), reflecting interpersonal trust and healthcare systems more generally (“I know that if my child was very ill, a doctor would come quickly” and “I don’t have any confidence in the NHS”), indicating system-level trust. Evidence of the association between trust in doctors and healthcare access at the 6-year timepoint was strongest for “the doctor in the clinic is always helpful”, reflecting how people may require interpersonal trust over system-level trust, to want to access healthcare.

These findings are consistent with previous literature highlighting the impact of interpersonal and system-level trust on help-seeking behaviours. Consistent with our results, Ewald and colleagues (32) found that expressing trust in healthcare professionals was associated with significantly lower odds of experiencing delayed healthcare, with the strongest associations being observed in the United Kingdom, compared to 20 other countries. Cavanagh and colleagues (74) also found that trust in healthcare professionals facilitates help-seeking for those with depressive symptoms. Our findings at the 6-year timepoint where associations were strongest for the interpersonal trust item (“the doctor in the clinic was always helpful”) aligns with evidence that suggests that interpersonal trust is particularly important in encouraging healthcare access. More broadly, trust in the healthcare system has also been associated with patient satisfaction (75) and healthcare outcomes (76). Conversely, those who mistrust healthcare systems are more likely to delay or avoid seeking treatment (77) and not adhere to treatment (78). The results of this study suggests that further research should be done to understand the role interpersonal trust and system-level trust play on healthcare access and the relationship between these factors.

Our findings suggest that the association between trust in doctors and healthcare access may differ between mothers and partners. At 21 months, mothers demonstrated consistent evidence for an association between trust in doctors and healthcare access for anxiety, depression and menstrual problems. At 6 years, mothers showed evidence of this association for two items for anxiety and one item for depression, whereas partners showed evidence of this association for one item for both anxiety and depression. These findings build on previous work that suggests that gender may impact the patient-physician relationship. However, in contrast to our findings, previous work suggests that women are less likely to trust their doctors compared to men (35,79,80). The associations between trust in doctors and healthcare access observed among mothers in this study may reflect the greater frequency of healthcare interactions during early parenthood which is likely to affect help-seeking. Though women consult healthcare providers more frequently than men (81), studies suggest that they face several challenges when attempting to access healthcare and receive treatment. Women often report feeling unheard or dismissed by practitioners (82) which is especially noticeable for women seeking care for menstrual disorders often feel like their pain is trivialised or completely disregarded (83). Therefore, while women may have greater engagement with healthcare services during parenthood, trust remains an important factor influencing whether they feel confident seeking care, particularly for conditions where they have historically experienced dismissal or inadequate recognition. Taken together, these findings suggest that the relationship between trust in doctors and healthcare access may differ by gender during early parenthood, highlighting the need to understand the unique barriers that parents may face when attempting to access healthcare.

Though the landscape of the healthcare system in the United Kingdom has shifted significantly since the data collection for this study, the results remain relevant. Disruptions and changes to the healthcare system have impacted the way in which people can access healthcare and how trusting they are of healthcare professionals. Studies have reported that trust in healthcare professionals has decreased over the course of the COVID-19 pandemic (84,85). This is compounded by the increased demand for primary care services, driven by recruitment crises in general practice, an ageing population and changes to care delivery since the COVID-19 pandemic (86). Following the pandemic, many GP practices have moved to telehealth, which has considerably affected patient-physician interactions, reducing face-to-face appointments. Though researchers have not reached a consensus on whether telehealth builds or neglects patient trust (87), it is important to recognise that this shift in the patient-physician relationship will change how physicians interact with their patients, which may affect the trust of patients. Several GP practices have also moved to a triage system, wherein patients who contact their practice will have to provide some information which is then screened (by nurses or digital technologies) to determine the type of consultation they receive and to manage appointment demand (88). This change has led to a decrease in the continuity of care, where patients do not see the same GP repeatedly (89). Repeated interactions with a healthcare provider have been shown to improve patient-physician trust (65) and as such, the move to a triage system will impact how confident patients are in their doctors’ abilities to provide them with appropriate consultations, diagnosis and treatment. The landscape of the current GP system sits in contrast with the system of the 1990s (when the data collection of this study took place) where wait times for appointments were lower, and patients would only see one GP for all their appointments, with greater trust in doctors to be expected. Therefore, results of this study suggest that as healthcare systems change, it remains pertinent to maintain trust in doctors to ensure people access the healthcare they need.

There are several limitations to this study that should be considered when interpreting these results. This study used cross-sectional data, making the results of this study unable to establish a causal relationship between trust in doctors and healthcare access for anxiety, depression and menstrual problems. This data may be susceptible to reverse causality in which accessing healthcare in the first place affects a patients’ trust in their doctor. Future research should explore whether there is a causal mechanism between trust and healthcare access and to explore the directionality of this relationship. The sample included in the present study is a relatively specific population, consisting of mothers and partners residing in the Avon region of the UK during the 1990s. As the sample was restricted to parents of young children, the findings may not be generalisable to individuals without children or to parents at different stages of the life course. Further, participants recruited into the ALSPAC cohort in the 1990s, were more likely than those in Britain to live in owner-occupied accommodation, have a higher socioeconomic position and be White (51). Analysis of this association should be conducted on a more diverse sample, to reflect how trust and healthcare experiences may differ across the life course and across differing groups such as different ethnic groups. The sample size of this study was also relatively small, especially for partners, leading to low statistical power which may not have detected small effect sizes. Further, partner data was not available for several covariates included in the study, leading to data from mothers being used as proxies for partner data. Participants were also not directly asked about how much they trusted their doctors; instead, exposure variables were chosen to represent trust. Believing that your doctor is “supportive”, “sympathetic”, “interested”, “helpful”, “easy to talk to” and “prepared to give time” is likely to result in an individual agreeing that they trust their doctor, but this is not assured. However, previous studies exploring how trust in physicians is developed generally agree that patients are believed to trust their doctors based on their competence, compassion, communication, dependability, reliability and confidentiality (24), which are covered by the exposure variables chosen for this study.

Overall, the results of this study highlight the importance of considering trust when understanding help-seeking behaviours for depression, anxiety and menstrual problems. Understanding the factors relating to healthcare access for anxiety, depression and menstrual problems is especially important due to their significant treatment gaps, preventing people with symptoms of these conditions from accessing the healthcare they need. Timely diagnosis and treatment are dependent on people accessing healthcare in the first instance. This work highlights the importance of maintaining the patient-physician relationship, even as healthcare systems evolve over time.

## Data Availability

Full instructions for applying for data access can be found here: http://www.bristol.ac.uk/alspac/researchers/access/. The ALSPAC study website contains details of all available data (http://www.bristol.ac.uk/alspac/researchers/our-data/)

https://github.com/diya-hoque/ALPSAC-trust-in-doctors

## Ethics statement

Ethical approval for the study was obtained from the ALSPAC Ethics and Law Committee and the Local Research Ethics Committees. Informed consent for the use of all data collected was obtained from participants following the recommendations of the ALSPAC Ethics and Law Committee at the time. Participants can contact the study team at any time to retrospectively withdraw consent for their data to be used. Study participation is voluntary and during all data collection sweeps, information was provided on the intended use of data.

The informed consent obtained from ALSPAC (Avon Longitudinal Study of Parents and Children) participants does not allow the data to be made available through any third party maintained public repository. Supporting data are available from ALSPAC on request under the approved proposal number, B4723. Full instructions for applying for data access can be found here: http://www.bristol.ac.uk/alspac/researchers/access/. The ALSPAC study website contains details of all available data (http://www.bristol.ac.uk/alspac/researchers/our-data/)

## Author Information

### Author Affiliations

School of Psychology and Neuroscience, University of Bristol, 12a Priory Road, Bristol, BS5 1TU Diya Hoque, Robyn E Wootton, Claire M A Haworth, Kayleigh E Easey

### Author Contributions

DH, RW, CH, and KE all contributed to the conceptualisation of this study. DH analysed, interpreted and visualised the data. Study write up was undertaken by DH, reviewing and editing on the manuscript was done by DH, RW, and KE. All authors reviewed and approved the final version of the manuscript.

## Disclosures and declarations

### Availability of data

R scripts for the analysis of data used in this study are available on https://github.com/diya-hoque/ALPSAC-trust-in-doctors. The informed consent of participants in ALSPAC does not allow the data to be made freely available through any third party-maintained repository. However, data used for this study can be requested from the ALSPAC executive. Full data on this process can be found here: https://www.bristol.ac.uk/alspac/researchers/access/.

## Acknowledgments

We are extremely grateful to all the families who took part in this study, the midwives for their help in recruiting them, and the whole ALSPAC team, which includes data collection staff, data and administrations staff, technical managers and the technical staff with the Bristol Bioresource Laboratory, based within the University of Bristol. The authors also thank Paul Madley-Dowd from the Bristol Medical School at the University of Bristol for his expert input on this work.

