## Supplementary materials for "Exploring associations between trust in doctors and the likelihood of accessing healthcare for anxiety, depression and menstrual problems"

### Supplementary Note

For the first timepoint (age 21 months), confounders were measured as follows: home ownership status (owned, mortgaged, privately rented, publicly rented and other), parity (1, 2, 3, 4, 5, 6+), educational attainment (university degree, <university degree) and socioeconomic position (measured using the British Registrar General’s Scale (Leete and Fox, 1977) (professional/managerial and other) and religion (Christian, non-Christian or no faith) were measured during pregnancy. Evaluation of own health (fit and well, mostly well, often unwell and never well), recent doctor change (yes/no), social support score (derived variable of 10 questions including questions such as “I have no one to share my feelings with” with responses being “this is exactly how I feel”, “this is often how I feel”, “this is sometimes how I feel” and “I never feel this way”, with any missing values set to the mode), weighted life events score (derived variable of 12 questions, including questions such as “you were admitted to hospital”, with responses being “this affected me a lot”, “this moderately affected me”, “this mildly affected me” “this did not affect me at all” “this did not happen to me”, with any missing values set to 0 unless all items were missing), marital status (married or not) and age were all measured at 21 months after birth.

For the second timepoint (age 6 years), confounders were included: for mothers, home ownership status, parity, educational attainment, religion, social class, marital status, social support score and recent doctor change, using the same measures described above. Weighted life events score was measured at 47 months postpartum, and evaluation of own health and age were measured at 6 years, 1 month postpartum. For partners, confounders were measured as follows: educational attainment and religion were measured during pregnancy, social support score and weighted life events score were measured at 1 year and 9 months postpartum and marital status was measured at 5 years, 1 month postpartum. Partner’s age and evaluation of own health were measured at 6 years, 1 month postpartum. Data on parity, home ownership status and a recent doctor change were not collected at the timepoint of interest (or beforehand) with mother data being used as a proxy.

Auxiliary variables, variables included in the imputation model but not the analysis model, were also used to aid in predicting missing values (90). Earlier measures of variables included in the analysis model and variables related and highly correlated to variables in the analysis model (e.g. car ownership status was used as an auxiliary variable for home ownership status) served as auxiliary variables.

### Correlation Matrices

#### Figure S1. Correlation matrix showing the correlations of the variables that were combined into one composite variable, reflecting trust in doctors, for mothers at timepoint 1

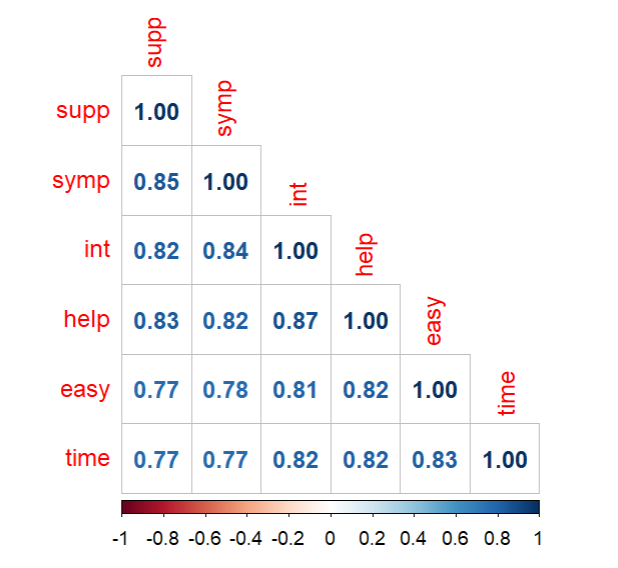

“Supp” = supportive; “symp” = sympathetic; “int” = interested; “help” = helpful; “easy” = easy to talk to; “time” = willing to give time

#### Figure S2. Correlation matrix showing the correlations of the variables reflecting trust in doctors at timepoint 2 (6 years) that remained separated, for mothers

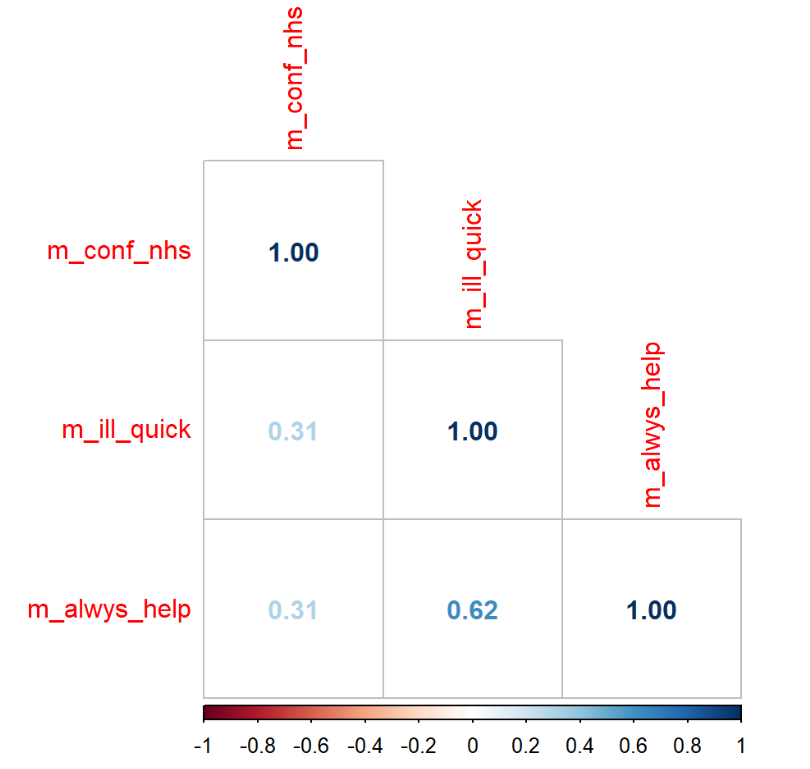

“m_conf_nhs” = I have no confidence in the national health service; “m_ill_quick” = I know that if my child was very ill, a doctor would come quickly; “m_alwys_help” = “the doctor in the clinic is always helpful”

#### Figure S3. Correlation matrix showing the correlations of the variables reflecting trust in doctors at timepoint 2 (6 years) that remained separated, for partners

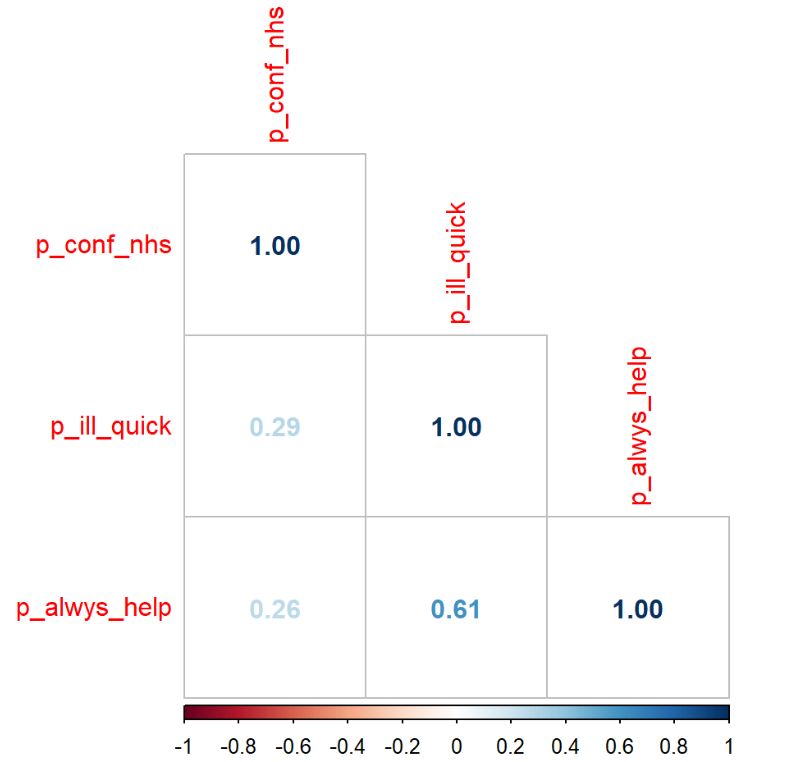

“p_conf_nhs” = I have no confidence in the national health service; “p_ill_quick” = I know that if my child was very ill, a doctor would come quickly; “p_alwys_help” = “the doctor in the clinic is always helpful”

Differences in correlation scores between variables at 21 months and 6 years may reflect differences in the focus of each question. At 21 months, variables are specifically and only asking participants about their relationship with their doctor, relating to interpersonal trust. In contrast, variables at 6 years asked participants about their opinions on their own doctor (“the doctor in the clinic is always helpful”), reflecting interpersonal trust and healthcare systems more generally (“I know that if my child was very ill, a doctor would come quickly” and “I don’t have any confidence in the NHS”), indicating system-level trust.

### Contingency Tables

#### Table S1. Participant characteristics of study variables by percentages, 21 months

|  | Anxiety | | | Depression | | | Menstrual problems | | |
| --- | --- | --- | --- | --- | --- | --- | --- | --- | --- |
| Patient Characteristics | **No** | **Yes, and saw Dr** | **Yes, but saw no Dr** | **No** | **Yes, and saw Dr** | **Yes, but saw no Dr** | **No** | **Yes, and saw Dr** | **Yes, but saw no Dr** |
| **Attitudes towards doctors** |  |  |  |  |  |  |  |  |  |
| 4-9 | 3% | 2% | 5% | 3% | 2% | 5% | 2% | 3% | 5% |
| 10-14 | 13% | 11% | 16% | 12% | 12% | 18% | 13% | 14% | 18% |
| 15-19 | 32% | 28% | 38% | 33% | 29% | 36% | 33% | 32% | 33% |
| 19-24 | 52% | 59% | 41% | 52% | 57% | 41% | 52% | 51% | 44% |
| **Age** |  |  |  |  |  |  |  |  |  |
| 15-25 | 17% | 15% | 15% | 16% | 22% | 20% | 16% | 22% | 16% |
| 26-30 | 40% | 36% | 36% | 40% | 37% | 37% | 39% | 39% | 36% |
| 31-35 | 31% | 36% | 34% | 32% | 31% | 31% | 32% | 28% | 35% |
| 36-40 | 10% | 11% | 12% | 11% | 9% | 11% | 11% | 9.7% | 12% |
| 41+ | 2% | 2% | 3% | 1% | 1% | 1% | 2% | 1.3% | 1% |
| **Parity** |  |  |  |  |  |  |  |  |  |
| 0 | 47% | 35% | 41% | 47% | 37% | 41% | 47% | 36% | 36% |
| 1 | 35% | 40% | 37% | 35% | 39% | 37% | 35% | 38% | 37% |
| 2 | 14% | 18% | 15% | 13% | 17% | 15% | 13% | 17% | 18% |
| 3+ | 4% | 7% | 7% | 5% | 7.0% | 7% | 5% | 9% | 9% |
| **Social class** |  |  |  |  |  |  |  |  |  |
| Other | 40% | 32% | 40% | 41% | 30% | 37% | 40% | 33% | 38% |
| Professional/Managerial | 60% | 68% | 60% | 59% | 70% | 63% | 60% | 67% | 62% |
| **Education** |  |  |  |  |  |  |  |  |  |
| O-level, Vocational, CSE, GCSE or none | 61% | 69% | 60% | 60% | 69% | 65% | 60% | 69% | 64% |
| A-level | 24% | 22% | 25% | 25% | 23% | 21% | 25% | 22% | 22% |
| Degree | 14% | 8.8% | 16% | 15% | 7.8% | 14% | 15% | 9.5% | 14% |
| **Home ownership** |  |  |  |  |  |  |  |  |  |
| Other | 3.0% | 3.4% | 3.0% | 2.8% | 3.3% | 4.2% | 2.9% | 3.4% | 3.5% |
| Publicly rented | 11% | 18% | 14% | 9.8% | 21% | 17% | 11% | 18% | 16% |
| Privately rented | 6.0% | 4.9% | 7.9% | 6.0% | 7.4% | 7.1% | 6.4% | 5.6% | 6.2% |
| Mortgaged | 78% | 73% | 73% | 79% | 67% | 69% | 77% | 72% | 74% |
| Owned | 2.3% | 0.8% | 2.4% | 2.3% | 1.3% | 2.4% | 2.5% | 1.1% | 1.1% |
| **Marital status** |  |  |  |  |  |  |  |  |  |
| Not currently married | 20% | 23% | 27% | 19% | 29% | 28% | 20% | 26% | 24% |
| Married | 80% | 77% | 73% | 81% | 71% | 72% | 80% | 74% | 76% |
| **Mum's religion** |  |  |  |  |  |  |  |  |  |
| Christian | 82% | 79% | 78% | 82% | 79% | 76% | 81% | 81% | 82% |
| Non-Christian | 3.4% | 4.7% | 6.2% | 3.3% | 5.0% | 5.9% | 3.7% | 4.9% | 4.4% |
| None | 15% | 17% | 16% | 14% | 16% | 18% | 15% | 14% | 13% |
| **Evaluation of own health** |  |  |  |  |  |  |  |  |  |
| Never well | 0.2% | 1.9% | 0.8% | 0.1% | 1.9% | 0.6% | 0.3% | 1.0% | 0.6% |
| Often unwell | 2.4% | 17% | 9.4% | 2.1% | 14% | 8.9% | 3.1% | 9.7% | 8.5% |
| Mostly well | 37% | 53% | 57% | 36% | 58% | 56% | 39% | 49% | 48% |
| Fit and well | 60% | 28% | 33% | 62% | 26% | 35% | 58% | 40% | 43% |
| **Recent dr change** |  |  |  |  |  |  |  |  |  |
| Recent Dr change | 16% | 20% | 21% | 16% | 20% | 21% | 17% | 18% | 14% |
| No recent Dr change | 84% | 80% | 79% | 84% | 80% | 79% | 83% | 82% | 86% |
| **Social support score** |  |  |  |  |  |  |  |  |  |
| 0-11 | 5.1% | 19% | 14% | 4.0% | 20% | 15% | 6.2% | 9.6% | 14% |
| 12-18 | 24% | 37% | 38% | 23% | 35% | 39% | 26% | 30% | 32% |
| 19-25 | 50% | 34% | 38% | 51% | 34% | 37% | 48% | 43% | 44% |
| 26-30 | 21% | 10% | 10% | 22% | 11% | 8.7% | 20% | 18% | 11% |
| **Weighted life events score** |  |  |  |  |  |  |  |  |  |
| 0-10 | 60% | 32% | 37% | 61% | 35% | 40% | 58% | 43% | 43% |
| 11-20 | 29% | 35% | 35% | 28% | 33% | 34% | 29% | 34% | 34% |
| 21-30 | 8.6% | 19% | 17% | 7.9% | 18% | 17% | 9.3% | 15% | 15% |
| 31-40 | 2.0% | 9.7% | 8.6% | 2.0% | 9.4% | 6.6% | 3.0% | 5.1% | 5.7% |
| 41-50 | 0.4% | 3.8% | 2.1% | 0.3% | 3.4% | 1.8% | 0.6% | 2.0% | 1.7% |
| 50+ | <0.1% | 1.3% | 0.8% | <0.1% | 1.2% | 0.5% | 0.2% | 0.6% | 0.8% |
| **Crown crisp anxiety score** |  |  |  |  |  |  |  |  |  |
| 0-4 | 75% | 19% | 26% | 76% | 27% | 36% | 68% | 51% | 50% |
| 5-8 | 20% | 31% | 43% | 21% | 29% | 36% | 23% | 29% | 31% |
| 9-12 | 3.9% | 29% | 23% | 3.3% | 26% | 20% | 7.3% | 13% | 12% |
| 13-16 | 0.7% | 21% | 7.9% | 0.4% | 17% | 6.8% | 2.1% | 7.2% | 7.0% |
| **Crown crisp depression score** |  |  |  |  |  |  |  |  |  |
| 0-4 | 80% | 38% | 48% | 83% | 37% | 47% | 74% | 61% | 59% |
| 5-8 | 18% | 35% | 39% | 16% | 37% | 40% | 21% | 29% | 31% |
| 9-12 | 2.5% | 25% | 12% | 1.4% | 23% | 12% | 4.6% | 9.1% | 8.6% |
| 13-16 | 0.1% | 2.5% | 1.3% | <0.1% | 2.4% | 1.3% | 0.3% | 1.2% | 1.0% |

#### Table S2. Participant characteristics of study variables for mothers and partners, who answered item 1, at 6 years

|  | Mothers | | | Partners | | |
| --- | --- | --- | --- | --- | --- | --- |
| Comparing Anxiety to confidence in NHS, with all covariates | **No** | **Yes, and saw Dr** | **Yes, but saw no Dr** | **No** | **Yes, and saw Dr** | **Yes, but saw no Dr** |
| **Home Ownership Status** |  |  |  |  |  |  |
| Publicly rented/Other | 5.9% | 7.5% | 6.6% | 5.6% | 3.4% | 5.5% |
| Privately rented | 3.4% | 4.9% | 3.9% | 2.9% | 4.2% | 3.2% |
| Mortgaged | 82% | 73% | 80% | 85% | 94 79% | 512 85% |
| Owned | 8.7% | 14% | 9.5% | 6.4% | 13% | 5.8% |
| **Parity** |  |  |  |  |  |  |
| 0 | 47% | 45% | 43% | 48% | 43% | 45% |
| 1 | 35% | 34% | 37% | 34% | 39% | 37% |
| 2 | 14% | 15% | 13% | 13% | 15% | 14% |
| 3+ | 4.4% | 5.7% | 6.9% | 4.5% | 2.5% | 3.9% |
| **Educational attainment** |  |  |  |  |  |  |
| O-level, Vocational, CSE, GCSE or none | 59% | 63% | 60% | 44% | 47% | 36% |
| A-level | 25% | 23% | 25% | 29% | 26% | 32% |
| Degree | 16% | 14% | 15% | 27% | 27% | 32% |
| **Religion** |  |  |  |  |  |  |
| None | 15% | 14% | 16% | 27% | 23% | 26% |
| Christian | 82% | 81% | 79% | 68% | 67% | 67% |
| Non-Christian | 3.6% | 5.3% | 4.7% | 4.8% | 9.3% | 7.7% |
| **Evaluation of own health** |  |  |  |  |  |  |
| Fit and well | 57% | 21% | 29% | 61% | 18% | 32% |
| Mostly well | 40% | 54% | 58% | 36% | 57% | 58% |
| Often unwell | 3.4% | 20% | 12% | 2.3% | 22% | 10% |
| Never well | 0.2% | 4.4% | 1.2% | 0.2% | 3.3% | 0.5% |
| **Social Support Score** |  |  |  |  |  |  |
| 0-11 | 4.8% | 12% | 9.5% | 3.7% | 9.8% | 7.4% |
| 12-18 | 24% | 34% | 34% | 38% | 35% | 49% |
| 19-25 | 51% | 42% | 44% | 55% | 52% | 40% |
| 26-30 | 21% | 13% | 12% | 3.9% | 3.3% | 3.6% |
| **Weighted life events score** |  |  |  |  |  |  |
| 0-9 | 53% | 32% | 35% | 61% | 42% | 42% |
| 10-19 | 33% | 38% | 37% | 30% | 48% | 37% |
| 20-29 | 11% | 20% | 20% | 7.5% | 7.8% | 17% |
| 30-38 | 3.5% | 9.9% | 7.2% | 1.3% | 2.2% | 4.4% |
| **Marital Status** |  |  |  |  |  |  |
| Not currently married | 8.5% | 16% | 8.5% | 7.4% | 8.8% | 11% |
| Married | 91% | 84% | 91% | 93% | 91% | 89% |
| **Recent doctor change** |  |  |  |  |  |  |
| Recent Dr change | 16% | 18% | 17% | 17% | 22% | 17% |
| No recent Dr change | 84% | 82% | 83% | 83% | 78% | 83% |
| **Age** |  |  |  |  |  |  |
| 15-30 | 16% | 17.9% | 14.1% | 6.5% | 8.0% | 4.2% |
| 31-40 | 72% | 70% | 72% | 68% | 60% | 61% |
| 41+ | 12% | 12% | 14% | 25% | 31% | 35% |
| **Socioeconomic Position** |  |  |  |  |  |  |
| Other | 59% | 59% | 62% | 46% | 48% | 40% |
| Professional/managerial | 41% | 41% | 38% | 54% | 52% | 60% |
| **No confidence in the NHS** |  |  |  |  |  |  |
| Never | 49% | 42% | 36% | 45% | 38% | 38% |
| Sometimes | 41% | 42% | 48% | 41% | 48% | 46% |
| Often | 8.4% | 11% | 13% | 10% | 10% | 12% |
| Exactly | 2.2% | 3.8% | 3.6% | 3.4% | 4.2% | 4.0% |

#### Table S3. Participant characteristics of study variables for mothers and partners, who answered item 2, at 6 years

|  | Mothers | | | Partners | | |
| --- | --- | --- | --- | --- | --- | --- |
| Comparing Anxiety to belief Dr would come quickly for sick child, with all covariates | **No** | **Yes, and saw Dr** | **Yes, but saw no Dr** | **No** | **Yes, and saw Dr** | **Yes, but saw no Dr** |
| **Home Ownership Status** |  |  |  |  |  |  |
| Publicly rented/Other | 5.9% | 7.5% | 6.6% | 5.6% | 3.4% | 5.5% |
| Privately rented | 3.4% | 4.9% | 3.9% | 2.9% | 4.2% | 3.2% |
| Mortgaged | 82% | 73% | 80% | 85% | 79% | 85% |
| Owned | 8.7% | 14% | 9.5% | 6.4% | 13% | 5.8% |
| **Parity** |  |  |  |  |  |  |
| 0 | 47% | 45% | 43% | 48% | 43% | 45% |
| 1 | 35% | 34% | 37% | 34% | 39% | 37% |
| 2 | 14% | 15% | 13% | 13% | 15% | 14% |
| 3+ | 4.4% | 5.7% | 6.9% | 4.5% | 2.5% | 3.9% |
| **Educational attainment** |  |  |  |  |  |  |
| O-level, Vocational, CSE, GCSE or none | 59% | 63% | 60% | 44% | 47% | 36% |
| A-level | 25% | 23% | 25% | 29% | 26% | 32% |
| Degree | 16% | 14% | 15% | 27% | 27% | 32% |
| **Religion** |  |  |  |  |  |  |
| None | 15% | 14% | 16% | 27% | 23% | 26% |
| Christian | 82% | 81% | 79% | 68% | 67% | 67% |
| Non-Christian | 3.6% | 5.3% | 4.7% | 4.8% | 9.3% | 7.7% |
| **Evaluation of own health** |  |  |  |  |  |  |
| Fit and well | 57% | 21% | 29% | 61% | 18% | 32% |
| Mostly well | 40% | 54% | 58% | 36% | 57% | 58% |
| Often unwell | 3.4% | 20% | 12% | 2.3% | 22% | 10% |
| Never well | 0.2% | 4.4% | 1.2% | 0.2% | 3.3% | 0.5% |
| **Social Support Score** |  |  |  |  |  |  |
| 0-11 | 4.8% | 12% | 9.5% | 3.7% | 9.8% | 7.4% |
| 12-18 | 24% | 34% | 34% | 38% | 35% | 49% |
| 19-25 | 51% | 42% | 44% | 55% | 52% | 40% |
| 26-30 | 21% | 13% | 12% | 3.9% | 3.3% | 3.6% |
| **Weighted life events score** |  |  |  |  |  |  |
| 0-9 | 53% | 32% | 35% | 61% | 42% | 42% |
| 10-19 | 33% | 38% | 37% | 30% | 48% | 37% |
| 20-29 | 11% | 20% | 20% | 7.5% | 7.8% | 17% |
| 30-38 | 3.5% | 9.9% | 7.2% | 1.3% | 2.2% | 4.4% |
| **Marital Status** |  |  |  |  |  |  |
| Not currently married | 8.5% | 16% | 8.5% | 7.4% | 8.8% | 11% |
| Married | 91% | 84% | 91% | 93% | 91% | 89% |
| **Recent doctor change** |  |  |  |  |  |  |
| Recent Dr change | 16% | 18% | 17% | 17% | 22% | 17% |
| No recent Dr change | 84% | 82% | 83% | 83% | 78% | 83% |
| **Age** |  |  |  |  |  |  |
| 15-30 | 16% | 17.9% | 14.1% | 6.5% | 8.0% | 4.2% |
| 31-40 | 72% | 70% | 72% | 68% | 60% | 61% |
| 41+ | 12% | 12% | 14% | 25% | 31% | 35% |
| **Socioeconomic Position** |  |  |  |  |  |  |
| Other | 59% | 59% | 62% | 46% | 48% | 40% |
| Professional/managerial | 41% | 41% | 38% | 54% | 52% | 60% |
| **If my child was very ill, a doctor would come quickly** |  |  |  |  |  |  |
| Exactly | 39% | 38% | 30% | 43% | 41% | 36% |
| Often | 27% | 26% | 30% | 30% | 33% | 35% |
| Sometimes | 26% | 25% | 30% | 21% | 22% | 23% |
| Never | 7.7% | 11% | 10% | 6.8% | 4.2% | 6.0% |

#### Table S4. Participants characteristics of study variables for mothers and partners, who answered item 3, at 6 years

|  | Mothers | | | Partners | | |
| --- | --- | --- | --- | --- | --- | --- |
| Comparing Anxiety to belief that the doctors in the clinic are always helpful, with all covariates | **No** | **Yes, and saw Dr** | **Yes, but saw no Dr** | **No** | **Yes, and saw Dr** | **Yes, but saw no Dr** |
| **Home Ownership Status** |  |  |  |  |  |  |
| Publicly rented/Other | 5.9% | 7.5% | 6.6% | 5.6% | 3.4% | 5.5% |
| Privately rented | 3.4% | 4.9% | 3.9% | 2.9% | 4.2% | 3.2% |
| Mortgaged | 82% | 73% | 80% | 85% | 79% | 85% |
| Owned | 8.7% | 14% | 9.5% | 6.4% | 13% | 5.8% |
| **Parity** |  |  |  |  |  |  |
| 0 | 47% | 45% | 43% | 48% | 43% | 45% |
| 1 | 35% | 34% | 37% | 34% | 39% | 37% |
| 2 | 14% | 15% | 13% | 13% | 15% | 14% |
| 3+ | 4.4% | 5.7% | 6.9% | 4.5% | 2.5% | 3.9% |
| **Educational attainment** |  |  |  |  |  |  |
| O-level, Vocational, CSE, GCSE or none | 59% | 63% | 60% | 44% | 47% | 36% |
| A-level | 25% | 23% | 25% | 29% | 26% | 32% |
| Degree | 16% | 14% | 15% | 27% | 27% | 32% |
| **Religion** |  |  |  |  |  |  |
| None | 15% | 14% | 16% | 27% | 23% | 26% |
| Christian | 82% | 81% | 79% | 68% | 67% | 67% |
| Non-Christian | 3.6% | 5.3% | 4.7% | 4.8% | 9.3% | 7.7% |
| **Evaluation of own health** |  |  |  |  |  |  |
| Fit and well | 57% | 21% | 29% | 61% | 18% | 32% |
| Mostly well | 40% | 54% | 58% | 36% | 57% | 58% |
| Often unwell | 3.4% | 20% | 12% | 2.3% | 22% | 10% |
| Never well | 0.2% | 4.4% | 1.2% | 0.2% | 3.3% | 0.5% |
| **Social Support Score** |  |  |  |  |  |  |
| 0-11 | 4.8% | 12% | 9.5% | 3.7% | 9.8% | 7.4% |
| 12-18 | 24% | 34% | 34% | 38% | 35% | 49% |
| 19-25 | 51% | 42% | 44% | 55% | 52% | 40% |
| 26-30 | 21% | 13% | 12% | 3.9% | 3.3% | 3.6% |
| **Weighted life events score** |  |  |  |  |  |  |
| 0-9 | 53% | 32% | 35% | 61% | 42% | 42% |
| 10-19 | 33% | 38% | 37% | 30% | 48% | 37% |
| 20-29 | 11% | 20% | 20% | 7.5% | 7.8% | 17% |
| 30-38 | 3.5% | 9.9% | 7.2% | 1.3% | 2.2% | 4.4% |
| **Marital Status** |  |  |  |  |  |  |
| Not currently married | 8.5% | 16% | 8.5% | 7.4% | 8.8% | 11% |
| Married | 91% | 84% | 91% | 93% | 91% | 89% |
| **Recent doctor change** |  |  |  |  |  |  |
| Recent Dr change | 16% | 18% | 17% | 17% | 22% | 17% |
| No recent Dr change | 84% | 82% | 83% | 83% | 78% | 83% |
| **Age** |  |  |  |  |  |  |
| 15-30 | 16% | 17.9% | 14.1% | 6.5% | 8.0% | 4.2% |
| 31-40 | 72% | 70% | 72% | 68% | 60% | 61% |
| 41+ | 12% | 12% | 14% | 25% | 31% | 35% |
| **The doctor in the clinic is always helpful** |  |  |  |  |  |  |
| Exactly | 40% | 40% | 30% | 40% | 48% | 32% |
| Often | 33% | 34% | 36% | 34% | 25% | 37% |
| Sometimes | 23% | 21% | 29% | 22% | 23% | 25% |
| Never | 3.7% | 4.5% | 4.6% | 4.3% | 4.2% | 5.6% |

#### Table S5. Participant characteristics of study variables for mothers and partners, who answered item 1, at 6 years

|  | Mothers | | | Partners | | |
| --- | --- | --- | --- | --- | --- | --- |
| Comparing Depression to confidence in NHS, with all covariates | **No** | **Yes, and saw Dr** | **Yes, but saw no Dr** | **No** | **Yes, and saw Dr** | **Yes, but saw no Dr** |
| **Home Ownership Status** |  |  |  |  |  |  |
| Publicly rented/Other | 5.6% | 9.8% | 6.4% | 5.3% | 2.9% | 8.2% |
| Privately rented | 3.3% | 5.0% | 4.3% | 2.8% | 4.3% | 4.0% |
| Mortgaged | 83% | 72% | 77% | 86% | 80% | 82% |
| Owned | 8.2% | 13% | 12% | 6.3% | 13% | 6.3% |
| **Parity** |  |  |  |  |  |  |
| 0 | 46% | 47% | 45% | 49% | 44% | 42% |
| 1 | 36% | 33% | 34% | 35% | 35% | 36% |
| 2 | 13% | 14% | 14% | 13% | 15% | 16% |
| 3+ | 4.3% | 5.6% | 7.7% | 4.2% | 5.1% | 6.0% |
| **Educational attainment** |  |  |  |  |  |  |
| O-level, Vocational, CSE, GCSE or none | 59% | 65% | 60% | 43% | 50% | 42% |
| A-level | 25% | 23% | 24% | 30% | 25% | 30% |
| Degree | 16% | 12% | 16% | 28% | 25% | 27% |
| **Religion** |  |  |  |  |  |  |
| None | 14% | 17% | 18% | 27% | 24% | 26% |
| Christian | 83% | 78% | 77% | 68% | 67% | 66% |
| Non-Christian | 3.4% | 5.2% | 5.6% | 5.0% | 8.7% | 7.9% |
| **Evaluation of own health** |  |  |  |  |  |  |
| Fit and well | 58% | 24% | 26% | 61% | 17% | 28% |
| Mostly well | 40% | 54% | 58% | 37% | 56% | 59% |
| Often unwell | 2.8% | 19% | 14% | 2.3% | 23% | 12% |
| Never well | 0.1% | 3.4% | 1.3% | 0.1% | 3.6% | 1.1% |
| **Social Support Score** |  |  |  |  |  |  |
| 0-11 | 4.2% | 13% | 11% | 3.6% | 9.1% | 11% |
| 12-18 | 23% | 34% | 38% | 38% | 35% | 51% |
| 19-25 | 52% | 40% | 40% | 54% | 55% | 35% |
| 26-30 | 21% | 14% | 11% | 4.1% | 0.9% | 2.9% |
| **Weighted life events score** |  |  |  |  |  |  |
| 0-9 | 53% | 33% | 35% | 61% | 38% | 39% |
| 10-19 | 32% | 38% | 37% | 30% | 45% | 37% |
| 20-29 | 11% | 21% | 20% | 7.4% | 11% | 20% |
| 30-38 | 3.5% | 8.3% | 7.8% | 1.4% | 5.6% | 4.2% |
| **Marital Status** |  |  |  |  |  |  |
| Not currently married | 8% | 13% | 10% | 7% | 11% | 12% |
| Married | 92% | 87% | 90% | 93% | 89% | 88% |
| **Recent doctor change** |  |  |  |  |  |  |
| Recent Dr change | 15% | 21% | 18% | 17% | 22% | 17% |
| No recent Dr change | 85% | 79% | 82% | 83% | 78% | 83% |
| **Age** |  |  |  |  |  |  |
| 15-30 | 14.9% | 22% | 15.7% | 6.2% | 9.2% | 4.6% |
| 31-40 | 73% | 70% | 69% | 68% | 59% | 63% |
| 41+ | 12% | 8.4% | 15% | 26% | 32% | 32% |
| **Socioeconomic Position** |  |  |  |  |  |  |
| Other | 59% | 63% | 60% | 45% | 53% | 43% |
| Professional/managerial | 41% | 37% | 40% | 55% | 47% | 57% |
| **No confidence in the NHS** |  |  |  |  |  |  |
| Never | 48% | 41% | 40% | 45% | 38% | 34% |
| Sometimes | 41% | 45% | 44% | 41% | 45% | 49% |
| Often | 8.8% | 10.0% | 11% | 10% | 11% | 12% |
| Exactly | 2.1% | 3.6% | 4.2% | 3.2% | 6.5% | 4.9% |

Table S6. Contingency table for mothers and partners, who answered item 2 and experienced depression, at 6 years

|  | Mothers | | | Partners | | |
| --- | --- | --- | --- | --- | --- | --- |
| Comparing Depression to belief Dr would come quickly for sick child, with all covariates | **No** | **Yes, and saw Dr** | **Yes, but saw no Dr** | **No** | **Yes, and saw Dr** | **Yes, but saw no Dr** |
| **Home Ownership Status** |  |  |  |  |  |  |
| Publicly rented/Other | 5.6% | 9.8% | 6.4% | 5.3% | 2.9% | 8.2% |
| Privately rented | 3.3% | 5.0% | 4.3% | 2.8% | 4.3% | 4.0% |
| Mortgaged | 83% | 72% | 77% | 86% | 80% | 82% |
| Owned | 8.2% | 13% | 12% | 6.3% | 13% | 6.3% |
| **Parity** |  |  |  |  |  |  |
| 0 | 46% | 47% | 45% | 49% | 44% | 42% |
| 1 | 36% | 33% | 34% | 35% | 35% | 36% |
| 2 | 13% | 14% | 14% | 13% | 15% | 16% |
| 3+ | 4.3% | 5.6% | 7.7% | 4.2% | 5.1% | 6.0% |
| **Educational attainment** |  |  |  |  |  |  |
| O-level, Vocational, CSE, GCSE or none | 59% | 65% | 60% | 43% | 50% | 42% |
| A-level | 25% | 23% | 24% | 30% | 25% | 30% |
| Degree | 16% | 12% | 16% | 28% | 25% | 27% |
| **Religion** |  |  |  |  |  |  |
| None | 14% | 17% | 18% | 27% | 24% | 26% |
| Christian | 83% | 78% | 77% | 68% | 67% | 66% |
| Non-Christian | 3.4% | 5.2% | 5.6% | 5.0% | 8.7% | 7.9% |
| **Evaluation of own health** |  |  |  |  |  |  |
| Fit and well | 58% | 24% | 26% | 61% | 17% | 28% |
| Mostly well | 40% | 54% | 58% | 37% | 56% | 59% |
| Often unwell | 2.8% | 19% | 14% | 2.3% | 23% | 12% |
| Never well | 0.1% | 3.4% | 1.3% | 0.1% | 3.6% | 1.1% |
| **Social Support Score** |  |  |  |  |  |  |
| 0-11 | 4.2% | 13% | 11% | 3.6% | 9.1% | 11% |
| 12-18 | 23% | 34% | 38% | 38% | 35% | 51% |
| 19-25 | 52% | 40% | 40% | 54% | 55% | 35% |
| 26-30 | 21% | 14% | 11% | 4.1% | 0.9% | 2.9% |
| **Weighted life events score** |  |  |  |  |  |  |
| 0-9 | 53% | 33% | 35% | 61% | 38% | 39% |
| 10-19 | 32% | 38% | 37% | 30% | 45% | 37% |
| 20-29 | 11% | 21% | 20% | 7.4% | 11% | 20% |
| 30-38 | 3.5% | 8.3% | 7.8% | 1.4% | 5.6% | 4.2% |
| **Marital Status** |  |  |  |  |  |  |
| Not currently married | 8% | 13% | 10% | 7% | 11% | 12% |
| Married | 92% | 87% | 90% | 93% | 89% | 88% |
| **Recent doctor change** |  |  |  |  |  |  |
| Recent Dr change | 15% | 21% | 18% | 17% | 22% | 17% |
| No recent Dr change | 85% | 79% | 82% | 83% | 78% | 83% |
| **Age** |  |  |  |  |  |  |
| 15-30 | 14.9% | 22% | 15.7% | 6.2% | 9.2% | 4.6% |
| 31-40 | 73% | 70% | 69% | 68% | 59% | 63% |
| 41+ | 12% | 8.4% | 15% | 26% | 32% | 32% |
| **Socioeconomic Position** |  |  |  |  |  |  |
| Other | 59% | 63% | 60% | 45% | 53% | 43% |
| Professional/managerial | 41% | 37% | 40% | 55% | 47% | 57% |
| **If my child was very ill, a doctor would come quickly** |  |  |  |  |  |  |
| Exactly | 39% | 36% | 32% | 43% | 41% | 32% |
| Often | 28% | 25% | 28% | 30% | 29% | 35% |
| Sometimes | 26% | 26% | 29% | 21% | 25% | 25% |
| Never | 7.3% | 13% | 11% | 6.5% | 5.8% | 7.7% |

#### Table S7. Participant characteristics of study variables for mothers and partners, who answered item 3, at 6 years

|  | Mothers | | | Partners | | |
| --- | --- | --- | --- | --- | --- | --- |
| Comparing Depression to belief that the doctors in the clinic are always helpful, with all covariates | **No** | **Yes, and saw Dr** | **Yes, but saw no Dr** | **No** | **Yes, and saw Dr** | **Yes, but saw no Dr** |
| **Home Ownership Status** |  |  |  |  |  |  |
| Publicly rented/Other | 5.6% | 9.8% | 6.4% | 5.6% | 3.4% | 5.5% |
| Privately rented | 3.3% | 5.0% | 4.3% | 2.9% | 4.2% | 3.2% |
| Mortgaged | 83% | 72% | 77% | 85% | 79% | 85% |
| Owned | 8.2% | 13% | 12% | 6.4% | 13% | 5.8% |
| **Parity** |  |  |  |  |  |  |
| 0 | 46% | 47% | 45% | 48% | 43% | 45% |
| 1 | 36% | 33% | 34% | 34% | 39% | 37% |
| 2 | 13% | 14% | 14% | 13% | 15% | 14% |
| 3+ | 4.3% | 5.6% | 7.7% | 4.5% | 2.5% | 3.9% |
| **Educational attainment** |  |  |  |  |  |  |
| O-level, Vocational, CSE, GCSE or none | 59% | 65% | 60% | 44% | 47% | 36% |
| A-level | 25% | 23% | 24% | 29% | 26% | 32% |
| Degree | 16% | 12% | 16% | 27% | 27% | 32% |
| **Religion** |  |  |  |  |  |  |
| None | 14% | 17% | 18% | 27% | 23% | 26% |
| Christian | 83% | 78% | 77% | 68% | 67% | 67% |
| Non-Christian | 3.4% | 5.2% | 5.6% | 4.8% | 9.3% | 7.7% |
| **Evaluation of own health** |  |  |  |  |  |  |
| Fit and well | 58% | 24% | 26% | 61% | 18% | 32% |
| Mostly well | 40% | 54% | 58% | 36% | 57% | 58% |
| Often unwell | 2.8% | 19% | 14% | 2.3% | 22% | 10% |
| Never well | 0.1% | 3.4% | 1.3% | 0.2% | 3.3% | 0.5% |
| **Social Support Score** |  |  |  |  |  |  |
| 0-11 | 4.2% | 13% | 11% | 3.7% | 9.8% | 7.4% |
| 12-18 | 23% | 34% | 38% | 38% | 35% | 49% |
| 19-25 | 52% | 40% | 40% | 55% | 52% | 40% |
| 26-30 | 21% | 14% | 11% | 3.9% | 3.3% | 3.6% |
| **Weighted life events score** |  |  |  |  |  |  |
| 0-9 | 53% | 33% | 35% | 61% | 42% | 42% |
| 10-19 | 32% | 38% | 37% | 30% | 48% | 37% |
| 20-29 | 11% | 21% | 20% | 7.5% | 7.8% | 17% |
| 30-38 | 3.5% | 8.3% | 7.8% | 1.3% | 2.2% | 4.4% |
| **Marital Status** |  |  |  |  |  |  |
| Not currently married | 8% | 13% | 10% | 7.4% | 8.8% | 11% |
| Married | 92% | 87% | 90% | 93% | 91% | 89% |
| **Recent doctor change** |  |  |  |  |  |  |
| Recent Dr change | 15% | 21% | 18% | 17% | 22% | 17% |
| No recent Dr change | 85% | 79% | 82% | 83% | 78% | 83% |
| **Age** |  |  |  |  |  |  |
| 15-30 | 14.9% | 22% | 15.7% | 5.6% | 8.0% | 4.2% |
| 31-40 | 73% | 70% | 69% | 68% | 60% | 61% |
| 41+ | 12% | 8.4% | 15% | 25% | 31% | 35% |
| **Socioeconomic Position** |  |  |  |  |  |  |
| Other | 59% | 63% | 60% | 46% | 48% | 40% |
| Professional/managerial | 41% | 37% | 40% | 54% | 52% | 60% |
| **The doctor in the clinic is always helpful** |  |  |  |  |  |  |
| Exactly | 40% | 38% | 32% | 40% | 48% | 32% |
| Often | 34% | 34% | 32% | 34% | 25% | 37% |
| Sometimes | 23% | 23% | 30% | 22% | 23% | 25% |
| Never | 3.5% | 4.7% | 5.8% | 4.3% | 4.2% | 5.6% |

### Regression analyses – 21 months

#### Table S8. Unadjusted and adjusted models exploring associations between trust in doctors and healthcare access for anxiety, depression and menstrual problems at 21 months, using imputed data

| **Item** | **Condition** | **Model** | **N** | **OR** | **95% CI** | **p-value** |
| --- | --- | --- | --- | --- | --- | --- |
| Trust in doctors | Anxiety | Unadjusted^a^ | 1849 | 1.09 | 1.06 - 1.11 | <0.001 |
|  |  | 1^b^ |  | 1.11 | 1.08 - 1.13 | <0.001 |
|  |  | 2^c^ |  | 1.11 | 1.08 - 1.14 | <0.001 |
|  |  | 3^d^ |  | 1.11 | 1.08 - 1.14 | <0.001 |
|  | Depression | Unadjusted^a^ | 2411 | 1.08 | 1.06 - 1.10 | <0.001 |
|  |  | 1^b^ |  | 1.10 | 1.07 - 1.12 | <0.001 |
|  |  | 2^c^ |  | 1.10 | 1.06 - 1.11 | <0.001 |
|  |  | 3^d^ |  | 1.10 | 1.07 - 1.12 | <0.001 |
|  | Menstrual problems | Unadjusted^a^ | 1723 | 1.04 | 1.02 - 1.06 | <0.001 |
|  |  | 1^b^ |  | 1.04 | 1.02 - 1.06 | <0.001 |
|  |  | 2^c^ |  | 1.04 | 1.02 - 1.07 | 0.002 |
|  |  | 3^d^ |  | 1.04 | 1.01 - 1.06 | 0.003 |

***^a^*** Unadjusted model

***^b^*** Model 1 (Baseline demographics). Adjusted for: age, parity, marital status, religion, socioeconomic position, educational attainment, home ownership status, crown crisp anxiety score and crown crisp depression score

***^c^*** Model 2 (Health experiences). Additionally adjusted for: self-rated health and recent doctor change

***^d^*** Model 3 (Social networks). Additionally adjusted for: social support score and weighted life events score

#### Table S9. Unadjusted and adjusted models exploring associations between trust in doctors and healthcare access for anxiety, depression and menstrual problems at 21 months, using original data

| **Item** | **Condition** | **Model** | **N** | **OR** | **95% CI** | **p-value** |
| --- | --- | --- | --- | --- | --- | --- |
| Sum of attitudes | Anxiety | Unadjusted | 1810 | 1.09 | 1.06 - 1.12 | <0.001 |
|  |  | 1 | 1260 | 1.11 | 1.08 - 1.15 | <0.001 |
|  |  | 2 | 1255 | 1.11 | 1.08 - 1.15 | <0.001 |
|  |  | 3 | 1253 | 1.11 | 1.08 - 1.15 | <0.001 |
|  | Depression | Unadjusted | 2360 | 1.08 | 1.06 - 1.10 | <0.001 |
|  |  | 1 | 1595 | 1.09 | 1.06 - 1.11 | <0.001 |
|  |  | 2 | 1583 | 1.09 | 1.06 - 1.12 | <0.001 |
|  |  | 3 | 1579 | 1.09 | 1.06 - 1.11 | <0.001 |
|  | Menstrual problems | Unadjusted | 1703 | 1.04 | 1.02 - 1.06 | <0.001 |
|  |  | 1 | 1122 | 1.03 | 1.01 - 1.06 | 0.012 |
|  |  | 2 | 1110 | 1.04 | 1.01 - 1.07 | 0.005 |
|  |  | 3 | 1106 | 1.03 | 1.01 - 1.06 | 0.017 |

***^a^*** Unadjusted model

***^b^*** Model 1 (Baseline demographics). Adjusted for: age, parity, marital status, religion, socioeconomic position, educational attainment, home ownership status, crown crisp anxiety score and crown crisp depression score

***^c^*** Model 2 (Health experiences). Additionally adjusted for: self-rated health and recent doctor change

***^d^*** Model 3 (Social networks). Additionally adjusted for: social support score and weighted life events score

#### Table S10. Unadjusted and adjusted models exploring associations between trust in doctors and healthcare access for anxiety, depression and menstrual problems at 21 months, using complete case data

| **Item** | **Condition** | **Model** | **N** | **OR** | **95% CI** | **p-value** |
| --- | --- | --- | --- | --- | --- | --- |
| Sum of attitudes | Anxiety | Unadjusted | 1253 | 1.09 | 1.06 - 1.13 | <0.001 |
|  |  | 1 |  | 1.10 | 1.06 - 1.13 | <0.001 |
|  |  | 2 |  | 1.10 | 1.07 - 1.13 | <0.001 |
|  |  | 3 |  | 1.09 | 1.06 - 1.13 | <0.001 |
|  | Depression | Unadjusted | 1579 | 1.08 | 1.05 - 1.10 | <0.001 |
|  |  | 1 |  | 1.08 | 1.05 - 1.11 | <0.001 |
|  |  | 2 |  | 1.08 | 1.06 - 1.11 | <0.001 |
|  |  | 3 |  | 1.07 | 1.05 - 1.10 | <0.001 |
|  | Menstrual problems | Unadjusted | 1106 | 1.03 | 1.01 - 1.06 | 0.014 |
|  |  | 1 |  | 1.04 | 1.01 - 1.07 | 0.005 |
|  |  | 2 |  | 1.04 | 1.02 - 1.07 | 0.002 |
|  |  | 3 |  | 1.03 | 1.00- 1.06 | 0.032 |

***^a^*** Unadjusted model

***^b^*** Model 1 (Baseline demographics). Adjusted for: age, parity, marital status, religion, socioeconomic position, educational attainment, home ownership status, crown crisp anxiety score and crown crisp depression score

***^c^*** Model 2 (Health experiences). Additionally adjusted for: self-rated health and recent doctor change

***^d^*** Model 3 (Social networks). Additionally adjusted for: social support score and weighted life events score

### Regression analyses – 6 years

#### Table S11. Unadjusted and adjusted models exploring association between trust in doctors and healthcare access for anxiety, for mothers and partners at 6 years, using imputed data

| **Condition** | **Item** | **Group** | **Model** | **N** | **OR** | **95% CI** | **p-value** |
| --- | --- | --- | --- | --- | --- | --- | --- |
| Anxiety | "I don't have any confidence in the NHS" | Mother | Unadjusted^a^ | 1892 | 1.14 | 1. 00 1-.29 | 0.05 |
|  |  |  | 1^b^ |  | 1.11 | 0.96 - 1.27 | 0.20 |
|  |  |  | 2^c^ |  | 1.17 | 1.01 - 1.35 | 0.03 |
|  |  |  | 3^d^ |  | 1.16 | 1.00 - 1.34 | 0.05 |
|  |  | Partner | Unadjusted^a^ | 734 | 0.90 | 0.72 - 1.12 | 0.30 |
|  |  |  | 1^b^ |  | 0.90 | 0.69 - 1.18 | 0.50 |
|  |  |  | 2^c^ |  | 1.06 | 0.79 - 1.42 | 0.70 |
|  |  |  | 3^d^ |  | 1.07 | 0.79 - 1.44 | 0.70 |
|  | "I know that if my child was very ill, a doctor would come quickly" | Mother | Unadjusted^a^ | 1892 | 1.12 | 1.02 - 1.24 | 0.02 |
|  |  |  | 1^b^ |  | 1.19 | 1.06 - 1.32 | 0.00 |
|  |  |  | 2^c^ |  | 1.21 | 1.08 - 1.35 | <0.001 |
|  |  |  | 3^d^ |  | 1.20 | 1.07 - 1.35 | 0.00 |
|  |  | Partner | Unadjusted^a^ | 734 | 1.05 | 0.86 - 1.27 | 0.70 |
|  |  |  | 1^b^ |  | 1.03 | 0.82 - 1.30 | 0.80 |
|  |  |  | 2^c^ |  | 1.15 | 0.90 - 1.46 | 0.30 |
|  |  |  | 3^d^ |  | 1.16 | 0.90 - 1.48 | 0.30 |
|  | "The doctor in the clinic is always helpful” | Mother | Unadjusted^a^ | 1892 | 1.27 | 1.13 - 1.42 | <0.001 |
|  |  |  | 1^b^ |  | 1.30 | 1.15 - 1.47 | <0.001 |
|  |  |  | 2^c^ |  | 1.36 | 1.20 - 1.55 | <0.001 |
|  |  |  | 3^d^ |  | 1.35 | 1.19 - 1.54 | <0.001 |
|  |  | Partner | Unadjusted^a^ | 734 | 1.29 | 1.03 - 1.62 | 0.029 |
|  |  |  | 1^b^ |  | 1.36 | 1.02 - 1.81 | 0.034 |
|  |  |  | 2^c^ |  | 1.42 | 1.06 - 1.90 | 0.02 |
|  |  |  | 3^d^ |  | 1.40 | 1.02 - 1.91 | 0.04 |

***^a^*** Unadjusted model

***^b^*** Model 1 (Baseline demographics). Adjusted for: age, parity, marital status, religion, socioeconomic position, educational attainment, home ownership status, crown crisp anxiety score and crown crisp depression score

***^c^*** Model 2 (Health experiences). Additionally adjusted for: self-rated health and recent doctor change

***^d^*** Model 3 (Social networks). Additionally adjusted for: social support score and weighted life events score

#### Table S12. Unadjusted and adjusted models exploring association between trust in doctors and healthcare access for anxiety, for mothers and partners at 6 years, using original dataset

| **Condition** | **Item** | **Group** | **Model** | **N** | **OR** | **95% CI** | **p-value** |
| --- | --- | --- | --- | --- | --- | --- | --- |
| Anxiety | "I don't have any confidence in the NHS" | Mother | Unadjusted | 1855 | 1.14 | 1.00 - 1.29 | 0.047 |
|  |  |  | 1 | 1212 | 1.17 | 0.99 - 1.40 | 0.071 |
|  |  |  | 2 | 1149 | 1.21 | 1.01 - 1.44 | 0.035 |
|  |  |  | 3 | 1146 | 1.20 | 1.01 -1.44 | 0.039 |
|  |  | Partner | Unadjusted | 725 | 1.02 | 0.80 - 1.32 | 0.90 |
|  |  |  | 1 | 327 | 1.03 | 0.73 - 1.48 | 0.90 |
|  |  |  | 2 | 429 | 1.06 | 0.74 - 1.55 | 0.70 |
|  |  |  | 3 | 394 | 1.06 | 0.72 - 1.58 | 0.80 |
|  | "I know that if my child was very ill, a doctor would come quickly" | Mother | Unadjusted | 1866 | 1.13 | 1.02 - 1.25 | 0.017 |
|  |  |  | 1 | 1159 | 1.15 | 1.01 - 1.32 | 0.037 |
|  |  |  | 2 | 1154 | 1.17 | 1.02 - 1.34 | 0.024 |
|  |  |  | 3 | 1151 | 1.17 | 1.02 - 1.34 | 0.026 |
|  |  | Partner | Unadjusted | 724 | 1.13 | 0.91 - 1.42 | 0.30 |
|  |  |  | 1 | 442 | 1.24 | 0.91 - 1.70 | 0.20 |
|  |  |  | 2 | 428 | 1.25 | 0.92 - 1.72 | 0.20 |
|  |  |  | 3 | 393 | 1.28 | 0.92 - 1.82 | 0.20 |
|  | "The doctor in the clinic is always helpful” | Mother | Unadjusted | 1824 | 1.27 | 1.14 - 1.43 | <0.001 |
|  |  |  | 1 | 1132 | 1.34 | 1.15 - 1.57 | <0.001 |
|  |  |  | 2 | 1127 | 1.38 | 1.18 - 1.62 | <0.001 |
|  |  |  | 3 | 1124 | 1.39 | 1.19 - 1.63 | <0.001 |
|  |  | Partner | Unadjusted | 722 | 1.29 | 1.03 - 1.62 | 0.03 |
|  |  |  | 1 | 442 | 1.47 | 1.07 - 2.04 | 0.02 |
|  |  |  | 2 | 428 | 1.47 | 1.07 - 2.05 | 0.02 |
|  |  |  | 3 | 393 | 1.49 | 1.06 - 2.14 | 0.02 |

***^a^*** Unadjusted model

***^b^*** Model 1 (Baseline demographics). Adjusted for: age, parity, marital status, religion, socioeconomic position, educational attainment, home ownership status, crown crisp anxiety score and crown crisp depression score

***^c^*** Model 2 (Health experiences). Additionally adjusted for: self-rated health and recent doctor change

***^d^*** Model 3 (Social networks). Additionally adjusted for: social support score and weighted life events score

#### Table S13. Unadjusted and adjusted models exploring association between trust in doctors and healthcare access for anxiety, for mothers and partners at 6 years, using complete case dataset

| **Condition** | **Item** | **Group** | **Model** | **N** | **OR** | **95% CI** | **p-value** |
| --- | --- | --- | --- | --- | --- | --- | --- |
| Anxiety | "I don't have any confidence in the NHS" | Mother | Unadjusted | 1146 | 1.13 | 0.96 - 1.34 | 0.15 |
|  |  |  | 1 |  | 1.17 | 0.99 - 1.39 | 0.076 |
|  |  |  | 2 |  | 1.21 | 1.01 - 1.44 | 0.035 |
|  |  |  | 3 |  | 1.20 | 1.01 - 1.44 | 0.039 |
|  |  | Partner | Unadjusted | 394 | 0.84 | 0.59 - 1.22 | 0.40 |
|  |  |  | 1 |  | 0.95 | 0.66 - 1.40 | 0.80 |
|  |  |  | 2 |  | 1.06 | 0.74 - 1.55 | 0.70 |
|  |  |  | 3 |  | 1.06 | 0.72 - 1.58 | 0.80 |
|  | "I know that if my child was very ill, a doctor would come quickly" | Mother | Unadjusted | 1151 | 1.12 | 0.98 - 1.28 | 0.09 |
|  |  |  | 1 |  | 1.15 | 1.01 - 1.32 | 0.036 |
|  |  |  | 2 |  | 1.17 | 1.02 - .134 | 0.024 |
|  |  |  | 3 |  | 1.17 | 1.02 - 1.34 | 0.026 |
|  |  | Partner | Unadjusted | 393 | 1.12 | 0.83 - 1.54 | 0.50 |
|  |  |  | 1 |  | 1.25 | 0.91 - 1.75 | 0.20 |
|  |  |  | 2 |  | 1.25 | 0.92 1.72 | 0.20 |
|  |  |  | 3 |  | 1.28 | 0.92 - 1.82 | 0.20 |
|  | "The doctor in the clinic is always helpful” | Mother | Unadjusted | 1124 | 1.32 | 1.14 - 1.54 | <0.001 |
|  |  |  | 1 |  | 1.36 | 1.16 - 1.59 | <0.001 |
|  |  |  | 2 |  | 1.38 | 1.18 - 1.62 | <0.001 |
|  |  |  | 3 |  | 1.39 | 1.19 - 1.63 | <0.001 |
|  |  | Partner | Unadjusted | 393 | 1.34 | 0.97 - 1.88 | 0.082 |
|  |  |  | 1 |  | 1.47 | 1.05 - 2.09 | 0.026 |
|  |  |  | 2 |  | 1.47 | 1.07 - 2.05 | 0.020 |
|  |  |  | 3 |  | 1.49 | 1.06 - 2.14 | 0.024 |

***^a^*** Unadjusted model

***^b^*** Model 1 (Baseline demographics). Adjusted for: age, parity, marital status, religion, socioeconomic position, educational attainment, home ownership status, crown crisp anxiety score and crown crisp depression score

***^c^*** Model 2 (Health experiences). Additionally adjusted for: self-rated health and recent doctor change

***^d^*** Model 3 (Social networks). Additionally adjusted for: social support score and weighted life events score

#### Table S14. Unadjusted and adjusted models exploring association between trust in doctors and healthcare access for depression, for mothers and partners at 6 years, using imputed data

| **Condition** | **Item** | **Group** | **Model** | **N** | **OR** | **95% CI** | **p-value** |
| --- | --- | --- | --- | --- | --- | --- | --- |
| Depression | "I don't have any confidence in the NHS" | Mother | Unadjusted^a^ | 2005 | 1.06 | 0.94 - 1.19 | 0.30 |
|  |  |  | 1^b^ |  | 1.05 | 0.92 - 1.19 | 0.50 |
|  |  |  | 2^c^ |  | 1.09 | 0.96 - 1.24 | 0.20 |
|  |  |  | 3^d^ |  | 1.08 | 0.95 - 1.23 | 0.30 |
|  |  | Partner | Unadjusted^a^ | 580 | 1.02 | 0.81 - 1.30 | 0.80 |
|  |  |  | 1^b^ |  | 1.16 | 0.86 - 1.56 | 0.30 |
|  |  |  | 2^c^ |  | 1.27 | 0.92 - 1.74 | 0.14 |
|  |  |  | 3^d^ |  | 1.24 | 0.9 - 1.70 | 0.20 |
|  | "I know that if my child was very ill, a doctor would come quickly" | Mother | Unadjusted^a^ | 2005 | 1.03 | 0.94 - 1.12 | 0.60 |
|  |  |  | 1^b^ |  | 1.09 | 0.99 - 1.20 | 0.10 |
|  |  |  | 2^c^ |  | 1.10 | 1.00 - 1.21 | 0.06 |
|  |  |  | 3^d^ |  | 1.08 | 0.98 - 1,20 | 0.12 |
|  |  | Partner | Unadjusted^a^ | 580 | 1.29 | 1.03 - 1.62 | 0.029 |
|  |  |  | 1^b^ |  | 1.30 | 1.00 - 1.69 | 0.046 |
|  |  |  | 2^c^ |  | 1.35 | 1.03 - 1.77 | 0.028 |
|  |  |  | 3^d^ |  | 1.30 | 0.99 - 1.72 | 0.06 |
|  | "The doctor in the clinic is always helpful” | Mother | Unadjusted^a^ | 2005 | 1.21 | 1.09 - 1.34 | <0.001 |
|  |  |  | 1^b^ |  | 1.27 | 1.14 - 1.42 | <0.001 |
|  |  |  | 2^c^ |  | 1.32 | 1.18 - 1.48 | <0.001 |
|  |  |  | 3^d^ |  | 1.30 | 1.16 - 1.46 | <0.001 |
|  |  | Partner | Unadjusted^a^ | 580 | 1.36 | 1.09 - 1.69 | 0.006 |
|  |  |  | 1^b^ |  | 1.41 | 1.05 - 1.89 | 0.021 |
|  |  |  | 2^c^ |  | 1.38 | 1.02 - 1.87 | 0.035 |
|  |  |  | 3^d^ |  | 1.46 | 1.05 - 2.03 | 0.023 |

***^a^*** Unadjusted model

***^b^*** Model 1 (Baseline demographics). Adjusted for: age, parity, marital status, religion, socioeconomic position, educational attainment, home ownership status, crown crisp anxiety score and crown crisp depression score

***^c^*** Model 2 (Health experiences). Additionally adjusted for: self-rated health and recent doctor change

***^d^*** Model 3 (Social networks). Additionally adjusted for: social support score and weighted life events score

#### Table S15. Unadjusted and adjusted models exploring association between trust in doctors and healthcare access for depression, for mothers and partners at 6 years, using original dataset

| **Condition** | **Item** | **Group** | **Model** | **N** | **OR** | **95% CI** | **p-value** |
| --- | --- | --- | --- | --- | --- | --- | --- |
| Depression | "I don't have any confidence in the NHS" | Mother | Unadjusted | 1957 | 1.06 | 0.95 - 1.19 | 0.30 |
|  |  |  | 1 | 1285 | 1.00 | 0.86 - 1.17 | >0.9 |
|  |  |  | 2 | 1210 | 1.03 | 0.88 - 1.20 | 0.70 |
|  |  |  | 3 | 1206 | 1.03 | 0.88 - 1.20 | 0.70 |
|  |  | Partner | Unadjusted | 569 | 1.03 | 0.81 - 1.30 | 0.80 |
|  |  |  | 1 | 339 | 1.07 | 0.76 - 1.52 | 0.70 |
|  |  |  | 2 | 318 | 1.08 | 0.76 - 1.55 | 0.70 |
|  |  |  | 3 | 284 | 1.13 | 0.77 - 1.67 | 0.50 |
|  | "I know that if my child was very ill, a doctor would come quickly" | Mother | Unadjusted | 1960 | 1.03 | 0.94 1.12 | 0.50 |
|  |  |  | 1 | 1209 | 1.09 | 0.97 1.23 | 0.20 |
|  |  |  | 2 | 206 | 1.10 | 0.98 1.24 | 0.11 |
|  |  |  | 3 | 1202 | 1.08 | 0.96 - 1.22 | 0.20 |
|  |  | Partner | Unadjusted | 567 | 1.16 | 0.95 1.44 | 0.20 |
|  |  |  | 1 | 325 | 1.27 | 0.95 1.72 | 0.12 |
|  |  |  | 2 | 316 | 1.26 | 0.93 - 1.72 | 0.14 |
|  |  |  | 3 | 282 | 1.24 | 0.89 - 1.73 | 0.20 |
|  | "The doctor in the clinic is always helpful” | Mother | Unadjusted | 1929 | 1.22 | 1.10 - 1.35 | <0.001 |
|  |  |  | 1 | 1189 | 1.25 | 1.10 - 1.41 | <0.001 |
|  |  |  | 2 | 1186 | 1.28 | 1.12 - 1.47 | <0.001 |
|  |  |  | 3 | 1182 | 1.29 | 1.12 - 1.48 | <0.001 |
|  |  | Partner | Unadjusted | 567 | 1.37 | 1.11 - 1.71 | 0.005 |
|  |  |  | 1 | 327 | 1.45 | 1.08 - 1.97 | 0.016 |
|  |  |  | 2 | 318 | 1.39 | 1.02 - 1.90 | 0.038 |
|  |  |  | 3 | 284 | 1.48 | 1.06 - 2.08 | 0.023 |

***^a^*** Unadjusted model

***^b^*** Model 1 (Baseline demographics). Adjusted for: age, parity, marital status, religion, socioeconomic position, educational attainment, home ownership status, crown crisp anxiety score and crown crisp depression score

***^c^*** Model 2 (Health experiences). Additionally adjusted for: self-rated health and recent doctor change

***^d^*** Model 3 (Social networks). Additionally adjusted for: social support score and weighted life events score

#### Table S16. Unadjusted and adjusted models exploring association between trust in doctors and healthcare access for depression, for mothers and partners at 6 years, using complete case dataset

| **Item** | **Condition** | **Group** | **Model** | **N** | **OR** | **95% CI** | **p-value** |
| --- | --- | --- | --- | --- | --- | --- | --- |
| Depression | "I don't have any confidence in the NHS" | Mother | Unadjusted | 1206 | 0.98 | 0.84 - 1.14 | 0.80 |
|  |  |  | 1 |  | 1.01 | 0.87 - 1.18 | 0.90 |
|  |  |  | 2 |  | 1.03 | 0.88 - 1.20 | 0.70 |
|  |  |  | 3 |  | 1.03 | 0.88 - 1.20 | 0.70 |
|  |  | Partner | Unadjusted | 284 | 0.96 | 0.68 - 1.37 | 0.80 |
|  |  |  | 1 |  | 1.05 | 0.73 - 1.53 | 0.80 |
|  |  |  | 2 |  | 1.08 | 0.76 - 1.55 | 0.70 |
|  |  |  | 3 |  | 1.13 | 0.77 - 1.67 | 0.50 |
|  | "I know that if my child was very ill, a doctor would come quickly" | Mother | Unadjusted | 1202 | 1.04 | 0.93 - 1.17 | 0.50 |
|  |  |  | 1 |  | 1.09 | 0.97 - 1.23 | 0.20 |
|  |  |  | 2 |  | 1.10 | 0.98 - 1.24 | 0.11 |
|  |  |  | 3 |  | 1.08 | 0.96 - 1.22 | 0.20 |
|  |  | Partner | Unadjusted | 282 | 1.10 | 0.83 - 1.47 | 0.500 |
|  |  |  | 1 |  | 1.22 | 0.9 - 1.68 | 0.200 |
|  |  |  | 2 |  | 1.26 | 0.93 - 1.72 | 0.140 |
|  |  |  | 3 |  | 1.30 | 1.13 - 1.49 | <0.001 |
|  | "The doctor in the clinic is always helpful” | Mother | Unadjusted | 1182 | 1.25 | 1.10 - 1.43 | <0.001 |
|  |  |  | 1 |  | 1.30 | 1.14 - 1.49 | <0.001 |
|  |  |  | 2 |  | 1.31 | 1.14 - 1.50 | <0.001 |
|  |  |  | 3 |  | 1.30 | 1.13 - 1.49 | <0.001 |
|  |  | Partner | Unadjusted | 284 | 1.32 | 0.98 - 1.82 | 0.075 |
|  |  |  | 1 |  | 1.47 | 1.07 - 2.05 | 0.021 |
|  |  |  | 2 |  | 1.39 | 1.02 - 1.90 | 0.038 |
|  |  |  | 3 |  | 1.48 | 1.06 - 2.08 | 0.023 |

***^a^*** Unadjusted model

***^b^*** Model 1 (Baseline demographics). Adjusted for: age, parity, marital status, religion, socioeconomic position, educational attainment, home ownership status, crown crisp anxiety score and crown crisp depression score

***^c^*** Model 2 (Health experiences). Additionally adjusted for: self-rated health and recent doctor change

***^d^*** Model 3 (Social networks). Additionally adjusted for: social support score and weighted life events score
